# Oxidative Status of the Ultra-Processed Foods in the Western Diet

**DOI:** 10.1101/2023.07.31.23293404

**Authors:** Lisaura Maldonado-Pereira, Carlo Barnaba, Ilce Gabriela Medina-Meza

## Abstract

Ultra-processed foods (UPFs) and their nutritional value have become a trending topic in the scientific community because of their increasing demand, and their potentially adverse effects on human health. Besides the poor nutritional value attributed to UPFs, countless studies have also reported the presence of different dietary oxidized substances in these meals. DOxS are associated with several chronic diseases such as cardiometabolic diseases (CMD), cancer, diabetes, Parkinson’s, and Alzheimer’s disease. In this study, a database of DOxS and other dietary metabolites detected in 63 UPFs meals part of the Western diet is reported. Significant differences were found in DOxS and phytosterol contents between ready-to-eat and fast food (FF). Putative biomarkers were suggested for RTE (brassicasterol) and FF (7α-OH and 7β-OH), as well as for all 6 different food categories: dairy (brassicasterol), eggs & derivatives (stigmasterol and β-sitosterol), meat & poultry (7α-OH), seafood, baby food (β-sitosterol), and others (campesterol). Ideally, the use of dietary biomarkers could potentially help in the future to identify in an early stage the presence of different chronic diseases, and even, prevent their development. Nevertheless, an exposure assessment is critical to understand the exposure level of DOxS and their relationship with different chronic diseases.

## 1. Introduction

Metabolic disorders are worldwide linked to poor dietary habits where Ultra processed foods (UPFs) are emerging as a serious threat to public health.^1, 2^ UPFs are a risk factor for metabolic disorders, including but not limited to metabolic and cardiovascular disorders, neurodegeneration, and cancer.^3–5^ UPFs have also been correlated with eating addictive-behaviors because of uncontrollable patterns of seeking and bingeing fat and sugar intake from highly palatable foods, causing long-term changes in the brain.^6–8^ The adverse effect of long-term exposure to UPFs is linked to their high caloric content, compared to the lower amount of essential micronutrients, such as minerals and vitamins.^9^ UPFs have become the most convenient and accessible food option for developed countries reaching 57% of the American’s caloric consumption in 2018,^10^ where 52.9% of UPFs are mostly consumed away from home by US youth aged 2-19 years.^11, 12^

The USDA’s 2020-2025 Dietary Guidelines state that 60% of Americans have one or more preventable chronic diseases related to dietary patterns ^13^. In the last decade, the USDA dietary and nutritional guidelines have been highlighting the need for healthy habits; however, these dietary recommendations still need to be met by the population. The most popular UPFs consumed in the USA are fast-food (FF) including small, large, and non-chain restaurants, and ready-to-eat (RTE), previously processed products, packed for sale which require minimum preparation at home.^14–16^ FF and RTE are highly top preference among children and youth ^17, 18^ because of the flavor, attractive design and marketing.^19, 20^

While is true that UPFs undergo extensive changes during industrial transformation to preserve microbiological safety and stability^21^ including high temperatures, exposure to light, type of cooking, packaging conditions, aging and storage, a caveat is that UPFs embody both an unbalanced ingredient profile as they are mostly constructed from a narrow range of cheap, extracted, refined and fractionated ingredients and detrimental processing that promotes oxidation of sensible compounds such as lipids and cholesterol,^22–26^ where changes are difficult to discriminate. Thus, the need for a reliable biomarker of processing is needed.

Dietary oxysterols (DOxS) are oxidized compounds derived from cholesterol oxidation in food, ^27, 28^ which have been proposed as markers of thermal processing of food.^26, 29, 30^ We also reported the occurrence of DOxS on infant formulations and baby foods, identifying 7-Ketocholesterol as marker of spray dry in infant formulations. ^26^ DOxS are bioactive lipids known to exert cytotoxic, pro-inflammatory and pro-apoptotic properties.^27, 31–33^ The establishment of food processing biomarkers will help to distinguish the extent of processing beyond a qualitative classification. Thus, the systematic mapping of DOxS in the food system is needed to identify levels of food processing, especially in food products within the category of UPFs^10^ to better assessment of the actual effect of processing on the UPFs nutritional quality.

The aim of the study is to fingerprint – for the first time – the content of DOxS and other bioactive lipids on >50 fast food and ready-to-eat meals consumed in the United States. These efforts represent the first step for building a large database that will be widely available to consumers and industry. Importantly, the database allows us to search for markers of food ultra-processing and evaluate its impact on the nutritional quality of the Western Diet. We believe that the information presented in this study will enable information regarding unintentional processing-formed compounds which will help to understand the relationship between them and the WD with the purpose of ensuring food nutritional safety and health.

## 2. Materials and Methods

### 2.1. Materials, Chemicals and Reagents

Methanol was from Sigma-Aldrich Chloroform was obtained from Omni Solv (Burlington, MA), hexane was purchased from VWR BDH Chemicals (Batavia, IL), 1-butanol and potassium chloride (KCl) from J. T. Baker (Allentown, PA) and diethyl ether was purchased from Fisher Chemical (Pittsburgh, PA). Sodium sulfate anhydrous (Na_2_SO_4_), Sodium chloride,^34^ were also purchased from VWR BDH Chemicals. standards of 7α-hydroxycholesterol (7α-OH), 7β-hydroxycholesterol (7β-OH), 5,6α-epoxycholesterol (5,6α-epoxy), 5,6β-epoxycholesterol (5,6β-epoxy), triol, 7-ketocholesterol (7-keto) were purchased from Steraloids (Newport, RI), and purified by using aminopropyl^35^ cartridges (500 mg/ 3 mL) from Phenomenex (Torrance, CA).

### 2.2. UPFs Selection, Collection and Preparation

The UPFs were selected, and classified into two main groups: fast-foods (FF) and ready-to-eat according to our previous study.^36^ FF (n=23) were collected from retail stores, supermarkets, food chains, restaurants, and takeaway in the Greater Lansing area (Michigan, USA) between February 2018 and October 2019, covering 75% of the national market.^16, 17, 37^ The RTE (n=39) were selected based on the Total Dietary Study 2011-2017 inventory ^38, 39^. The complete list of the food meals, their respective test code, and their group is provided in **Table 1**. Additionally, foods were grouped into categories according to the fat source as follows: eggs and egg’s derivatives (E), dairy products (D), meat and poultry (MP), seafood (S), baby food (BF). Food items that did not fit in any of the previous categories, such as potato-products (potato crisps with and without added flavors, and French fries from restaurants and takeaway, pasta, salad dressings, and popcorn (sweet or salty) were grouped as other products (O).

**Table 1.** Ultra-processed foods meals ID used in this study, divided by group and category.

| Category | Sample ID | Ultra-processed Foods (UPF) | Group |
| --- | --- | --- | --- |
|  | D1 - RTE | American Cheese – Happy Farms | Ready to Eat (RTE) |
|  | D2 - RTE | Cheddar cheese – Happy Farms |  |
|  | D3 - RTE | Margarine (regular, not low-fat, salted) – Countryside Creamery |  |
|  | D5 - RTE | Cream (half & half) -Meijer |  |
|  | D6 - RTE | Swiss cheese - Kroger |  |
|  | D7 - RTE | Cream cheese – Happy Farms |  |
|  | D8 - RTE | Ice cream (regular, not low-fat, vanilla) - Purple Cow |  |
|  | D9 - RTE | Yogurt (low-fat, fruit flavored) - Yoplait |  |
|  | D10 - RTE | Chocolate milk - Nesquik |  |
|  | D11 - RTE | Infant formula – Little Journey (with iron milk-based powder) |  |
|  | * D4 – RTE | Butter – Praire Farms |  |
| **Meat & Poultry** | MP1 - RTE | Bologna - Eckrich |  |
|  | MP2 - RTE | Salami – Oscar Mayer |  |
|  | MP3 - RTE | Soup bean w/bacon/pork (canned, prepared w/water) - Campbell's |  |
|  | MP4 – RTE | Chili con carne w/beans (canned) - Campbell's |  |
|  | MP5 – RTE | Lasagna w/meat (frozen, heated) – Michael Angelo's |  |
|  | MP6 – RTE | Chicken noodle soup - Kroger |  |
|  | MP7 – RTE | Beef and vegetables soup – Kroger |  |
|  | MP8 – RTE | Mini Ravioli - Chef Boyardee |  |
|  | MP9 - RTE | Spaghetti - Chef Boyardee |  |
| **Seafood** | S1 - RTE | Clam chowder (New England, canned, prep w/ whole milk) – Kroger |  |
| **Eggs & derivatives** | E1 - RTE | Mayonnaise (regular, bottled) – Hellmann's |  |
|  | E2 - RTE | Macaroni salad (from grocery/deli) - Meijer |  |
| **Baby food** | BF1 - RTE | Baby food - beef and broth/gravy – Beech Nut |  |
|  | BF2 - RTE | Baby food – chicken and broth/gravy - Gerber |  |
|  | BF3 - RTE | Baby food - vegetables and beef - Gerber |  |
|  | BF4 - RTE | Baby food - vegetables and chicken - Gerber |  |
|  | BF5 - RTE | Baby food - chicken noodle dinner - Gerber |  |
|  | BF6 - RTE | Baby food - macaroni, tomato and cheese - Gerber |  |
|  | BF7 - RTE | Baby food - turkey and rice - Gerber |  |
|  | BF8 - RTE | Baby food – turkey and broth/gravy – Beech Nut |  |
|  | BF9 - RTE | Baby food – fruit yogurt - Gerber |  |
|  | BF10 - RTE | Baby food – chicken with rice - Gerber |  |
|  | BF11 - RTE | Baby food – vegetables and turkey - Gerber |  |
|  | BF12 - RTE | Baby food – macaroni and cheese with vegetables - Gerber |  |
|  | BF13 - RTE | Pasta pick-ups (cheese ravioli) - Gerber |  |
| **Others** | O1 - RTE | Popcorn w/butter (microwave) - Kroger |  |
|  | O2 - RTE | Salad dressing (creamy/buttermilk type, regular) – Aldi's Tuscan Garden |  |
|  | O3 - RTE | Macaroni & cheese (boiled) - Kraft |  |
|  | O4 - RTE | Macaroni & cheese (microwaved) - Kraft |  |
| **Meat & Poultry** | MP10 - FF | Hamburger on bun – McDonald's | **Fast Food (FF)** |
|  | MP11 - FF | Chicken nuggets – McDonald's |  |
|  | MP12 - FF | Cheeseburger on bun – McDonald's |  |
|  | MP13 - FF | Steak tacos w/beans, lettuce, rice and cheese - Chipotle |  |
|  | MP14 - FF | Cheese and chicken quesadilla - Chipotle |  |
|  | MP15 - FF | Chicken burrito w/lettuce, cheese, pico - Chipotle |  |
|  | MP16 - FF | Chicken drumstick - KFC |  |
|  | MP17 - FF | Chicken wing - KFC |  |
|  | MP18 - FF | Beef w/vegetables - Panda Express |  |
|  | MP19 - FF | Chicken w/vegetables - Panda Express |  |
|  | MP20 - FF | Chicken filet - (broiled sandwich) – Chick Fil'A |  |
|  | MP21 - FF | Roast beef, ham & Provolone - Jimmy Johns |  |
|  | MP22 - FF | Sliced turkey and bacon - Jimmy Johns |  |
|  | MP23 - FF | Supreme pizza - Marco's Pizza |  |
|  | MP24 - FF | Pepperoni pizza, hand tossed - Domino's |  |
| Seafood | S2 - FF | Fish sandwich on bun – McDonald's |  |
|  | S3 - FF | Fried Shrimp – Panda Express |  |
| Others | O5 - FF | French-Fries – McDonald's |  |
|  | O6 - FF | McDonald's Biscuit - Big Breakfast |  |
|  | O7 - FF | McDonald's Hotcakes - Big Breakfast |  |
|  | O8 - FF | Biscuit - KFC |  |
|  | O9 - FF | French Fries - KFC |  |
|  | O10 - FF | Mashed potato - KFC |  |

\* The only processed (category 3 NOVA) food in this study.

FF were purchased from the selected franchises and brought to the laboratory for immediate analysis. RTE meals were purchased from different local supermarket stores, and immediately brought to the laboratory. Storage conditions were followed according to the label instructions (fresh foods were kept in a fridge at 4°C and frozen meals were kept at −20°C or the temperature indicated in the label).

Once the UPF samples arrived at the laboratory, we promptly recorded the following essential information: (1) UPF’s name; (2) price; (3) place of purchase, date, and time of collection; (4) type of food (Ready-to-Eat - RTE or Fast-Food - FF); (5) nutritional declaration (including energy, fat, saturated fatty acids, carbohydrates, sugars, fiber, protein, and salt); (6) portion size; (7) list of ingredients; (8) expiration date; and (9) any other pertinent details. For certain UPFs that required additional preparation before analysis, we strictly followed the manufacturer’s instructions, which are listed in **Table 2**. We conducted the analysis using the Sensory Lab kitchen facility at Michigan State University, ensuring accuracy and precision. The experiments were performed with three biological replicates, and we stored the samples appropriately, taking into consideration their specific food matrices.

**Table 2.**
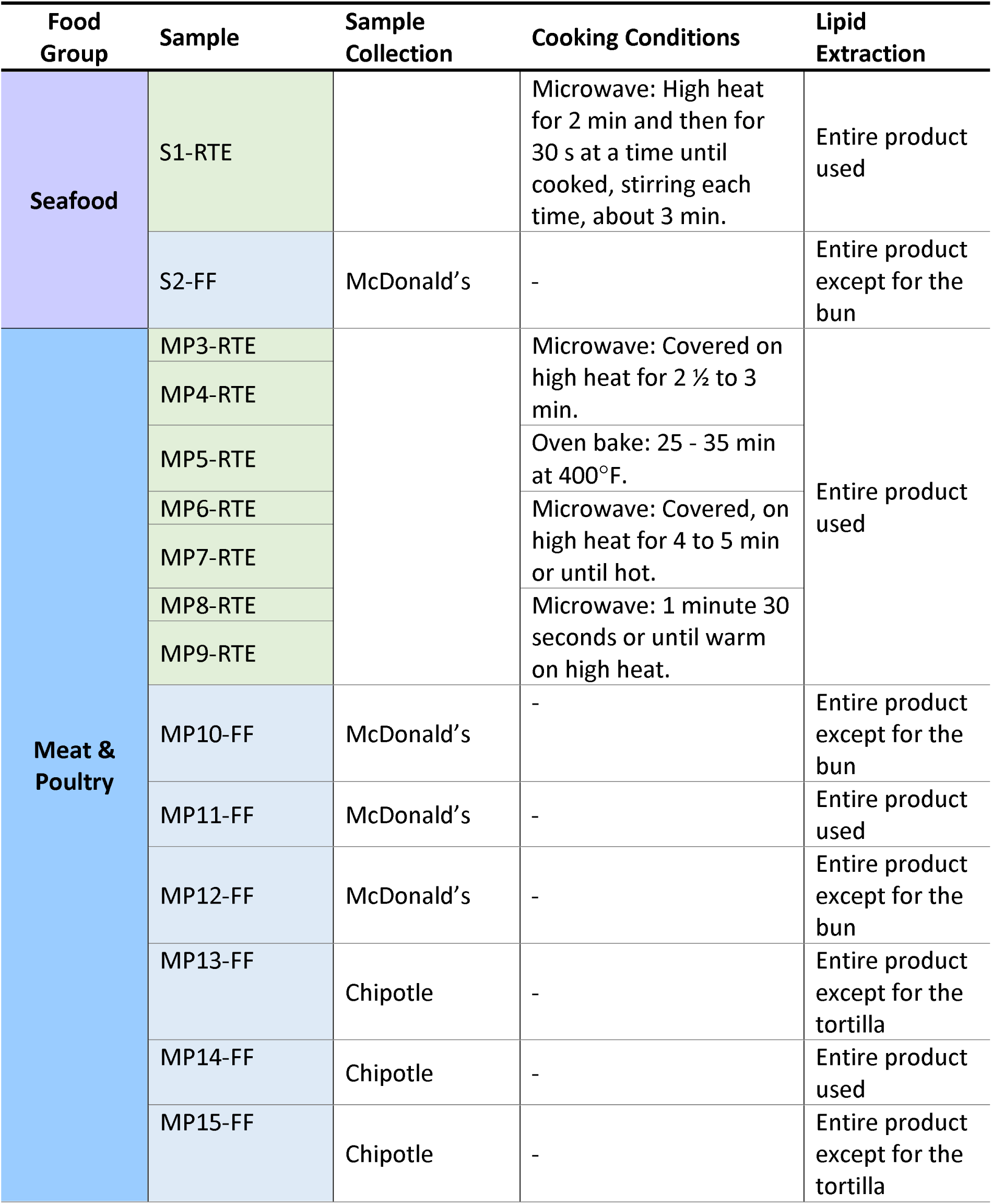
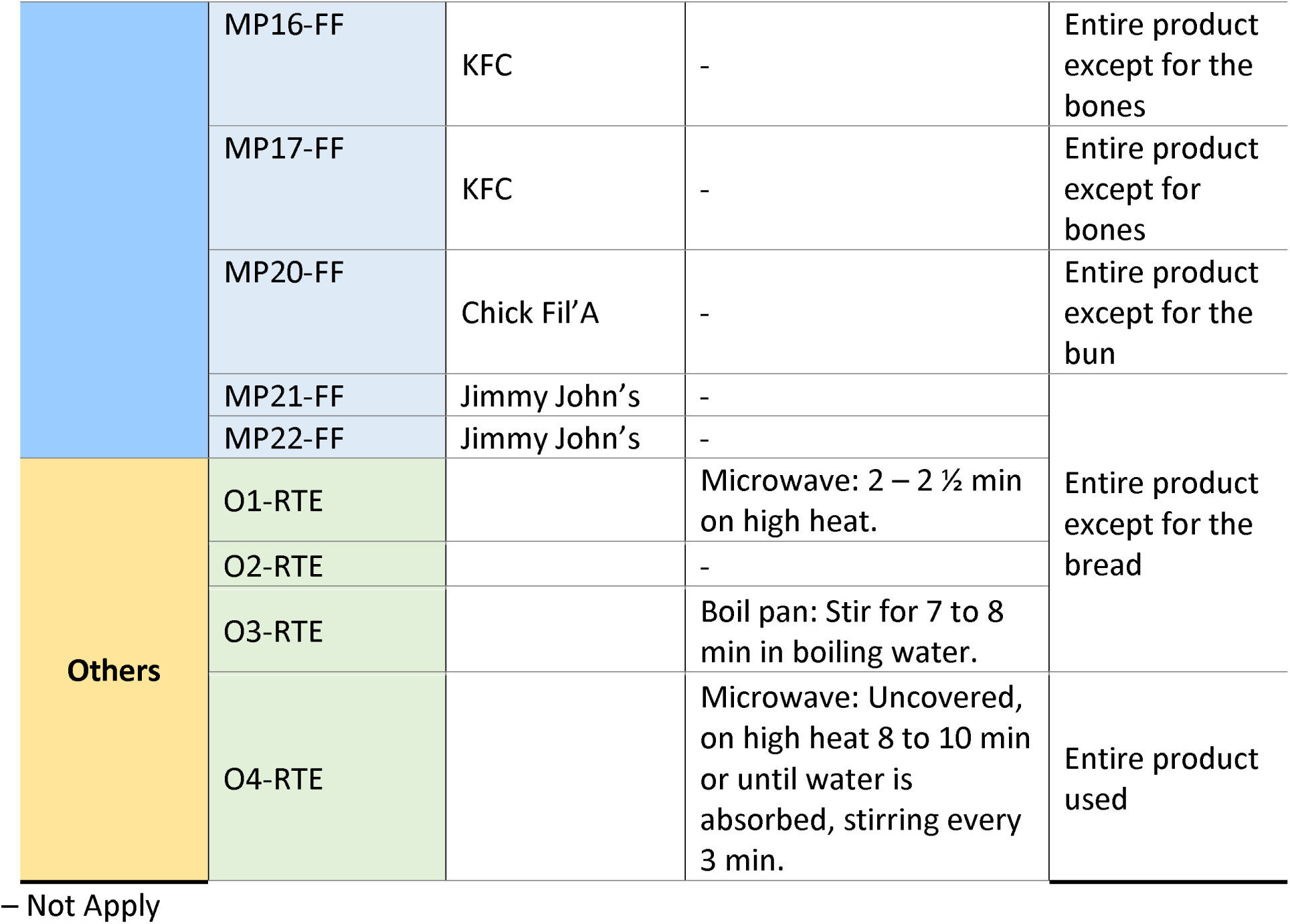
Additional cooking preparations according to the label’s instructions.

### 2.3. Lipid Extraction

A cold lipid extraction according to Folch and coworkers^40^ with some modifications depending on the food matrix was performed. Thirty grams of sample were homogenized using an Ultra-Turrax® (Tekmar TP 18/10S1 Cincinnati, OH) for 3 min at 300 rpm, Then, placed in a 500 mL glass bottle with screwcap with 200 mL of a chloroform:methanol solution (1:1, v/v). Bottle was kept in an oven at 60°C for 20 min before adding 100 mL chloroform. After 2 min of vortex, the content of the bottle was filtered. The filtrate was mixed thoroughly with a 100 mL of 1 M KCl solution. Samples were left overnight at 4°C. Then, the lower phase containing lipids was collected and dried at 60°C with a vacuum evaporator at 25 in Hg. Total fat content was determined gravimetrically and fatty acids profiles were previously reported.^36^ In addition, fat content is reported in both units—percentage of total fatty acid weight (% w/w) per 100 g of fresh sample and weight of total fat per serving size (g per serving size)—for nutritional comparison purposes.

### 2.4. Thiobarbituric Acid Reactive Substances (TBARS)

The method modified by Miller ^41^, was used to measure lipid oxidation in all UPFs with some modifications. Briefly, 60 mg of fat were weighed into a 10 mL glass test tube with screw cap and the following reagents were added: 100 μL BHT solution (0.2 mg/mL in water) and 4.9 mL extracting solution (10% TCA in 0.l M H_2_PO_3_). Sample blanks were analyzed along with each sample. A standard curve of 0-5 mL TEP solution (10 μM) was prepared. Test tubes were incubated overnight in the dark at room temperature. TBARS were expressed as μg malondialdehyde (MDA)/g fat sample.

### 2.5. Total Cholesterol, Tocopherols and Phytosterols Content

A simultaneous isolation method for cholesterol, tocopherols, and phytosterols developed in our laboratory^26^ was used. 200 mg of lipid were mixed with 50 μg of 19-hydroxycholesterol and 140 μg of 5α-cholestane as internal standards for the determination of phytosterols, tocopherols and total cholesterol, respectively. Subsequently, the sample was mixed with 10 mL of 1 N KOH solution in methanol and left it under gentle agitation and covered from the light for at least 15 h. One-tenth of the unsaponifiable matter was subjected to silylation.

Sample was mixed with 100 μL of pyridine and 100 μL of the Sweeley’s reactive mixture (pyridine/hexamethyldisilazane/trimethylchlorosilane, 10:2:1, v/v/v) at 75°C for 45 min, dried under nitrogen stream and dissolved in 1 mL of n-hexane. One microliter of the silylated solution was injected into a gas chromatograph using a fitted with a Zebron™ZB-5HT (Phenomenex, Torrance, CA) capillary column (30 m × 0.25 mm × 0.25 μm). Oven temperature conditions were set up as follow: 260 to 300°C at 2.5°C/min, then from 300 to 320°C at 8°C/min, and finally hold at 320°C for 1 min. The injector and detector were both set at 320°C. Helium was used as the carrier gas at a flow rate of 54.0 mL/min, the split ratio at 1:50, and pressure constant at 134 kPa.

### 2.6. DOxS Quantification

The other nine-tenths of the sample was used for the quantification of dietary DOxS. Purification by NH_2_-SPE was performed according to Kilvington et al. ^26^ The enriched fraction was recovered and evaporated to dryness under a nitrogen stream. Subsequently, the purified fraction was silylated with Sweeley’s reactive mixture (at 75 °C for 45 min), dried under a nitrogen stream and dissolved in 1 mL of n-hexane.

DOxS acquisition was done using a GCMS-QP2010 SE single quadrupole Shimadzu Corporation (Kyoto, Japan). with one μL of the silylated sample. DOxS were injected into a single quad GCMS-QP2010 SE with the following conditions: from 250 to 280 °C at 2°C/min, hold at 280 °C for 7 min, and from 280 to 315°C at 1.5 °C/min. Helium was used as a carrier gas (flow rate of 0.37 mL/min); the split ratio was 1:15 and pressure 49.2 KPa. The interface temperature for the GC-MS was 320 °C, with the electron multiplier voltage set at 0 kV and injector at 320 °C. A fused silica column (30 m x 0.25 mm i.d. x 0.25 um thickness) coated with 5% phenyl polysyloxane (Phenomenex Zebron ZB-5) and selected ion monitoring (SIM) mode were used.

### 2.7. Statistical Analysis

Descriptive statistics were calculated overall, by category, and by food origin. Both mean, and confidence interval (95%) were computed. When comparing RTE vs FF, data did not follow a normal distribution after a density plot analysis and was not homogenous after a residual plot analysis. Therefore, a Matt-Whitney *U*-test was performed, at *p* < 0.05 significance level. Statistical differences between food categories were evaluated by using the non-parametric Kruskal-Wallis ANOVA by Ranks test, at *p* < 0.05 significance level. Spearman’s correlation across nutritional and non-nutritional variables was also tested. All the statistical analyses were computed using Rstudio (Version 1.4.1717 © 2009-2021 RStudio, PBC.). Principal Component Analysis was performed in OriginPro (v. 2023, OriginLab) using correlation matrix on normalized data.

## 3. Results

### 3.1. Fat, sterols content, and FAME nutritional indexes in RTE and FF

We performed quantitative assessment of total fat, cholesterol and phytosterols content in the selected UPFs (**Figure 1**). Fat content in UPFs was statistically similar between FF and RTE when measured on a weight basis; however, if computed according to the serving size, FF showed a significantly higher amount of fat (**Fig. 1A**). As expected, fat-enriched food categories were dairy, and egg and derivatives, whereas seafood categories had the higher fat content when computed on serving basis (**Fig. 1B**).

**Figure 1.**
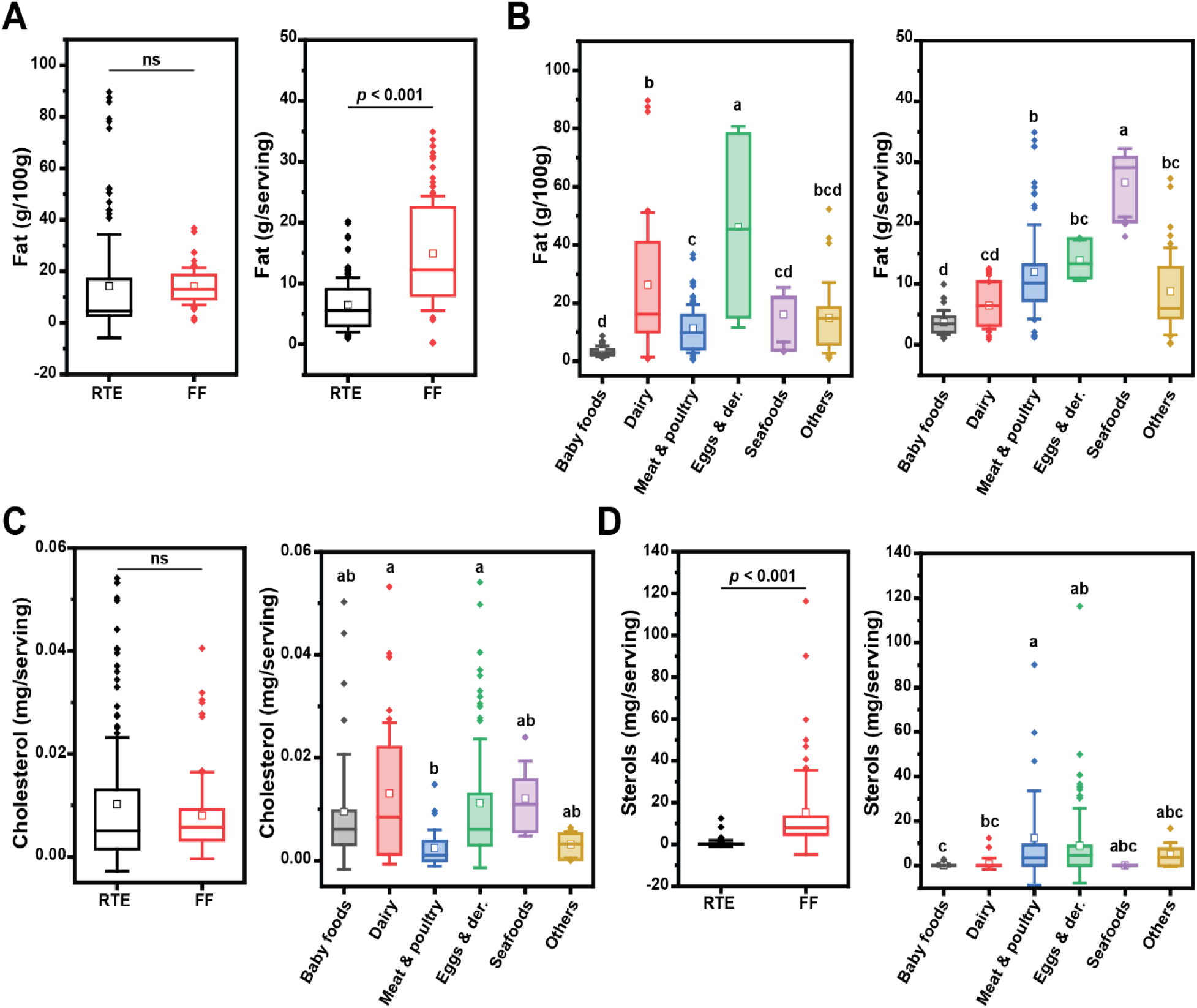
Total fat (A, B), cholesterol (C), and sterols (D) contents in UPFs by group and category. Box plots represent confidence interval (25-75%) ± 1SD, line indicates median and white square indicates mean; outliers are also represented as diamonds. When comparing RTE and FF a paired *t*-test was used. The grouping letters in the graph represent distinct categories within the variable under analysis, highlighting significant differences between the groups as determined by the ANOVA, followed by post-hoc Tukey test (p<0.05).

Cholesterol and phytosterols are the sterol-like compounds present in UPFs from animal and vegetable origin, respectively. For cholesterol, no significant difference between RTE and FF was observed, and only marginal differences between food categories were observed (**Fig. 1C**). FF had a higher sterols content (**Fig. 1D**); when looking at food categories, meat and egg products showed higher variance in sterols content (**Fig. 1D**, right panel). As expected, cholesterol was abundant in dairy products and eggs products (category D and E, respectively). Dietary cholesterol from dairy products ranges between 80 mg to 110 g ^42^. The lack of vegetable oils and other ingredients source of phytosterols, drive the Dairy category to rank 4^th^ in the phytosterols content. Interestingly, Dairy products also were 4^th^ in the DOxS content. Eggs and derivatives category had the highest phytosterols content among all categories and was the 2^nd^ highest category in cholesterol content (**Fig. 1 C,D**). The UPFs comprising the Eggs category, Mayonnaise (E1-RTE) and macaroni salad (E2-RTE) are not solely composed of eggs. They contain a significant amount of soybean oil ^43, 44^ which is a main source of phytosterols and places this category in the first place compared to the rest of the categories. E’s position in the cholesterol content assay was expected due to its already known high cholesterol content which ranges from 193-275 mg per egg ^45–48^. BF have high cholesterol content and a discrete amount of phytosterol (**Fig. 1C**), due to their formulation that includes meat-derivatives as main ingredients, as well as vegetable oils ^26, 49–51^. Meat and Poultry had lower cholesterol content than Dairy and eggs category (**Fig. 1C**), with a few exceptions. MP6-RTE, MP16-FF, MP17-FF, and BF1-RTE (samples principally made of chicken, and beef) contain the highest cholesterol amounts (**Table S2, Supplemental Information**). Therefore, the ingredients used during the confection of these meals need to be taken into consideration. The noodles present in the chicken noodle soup (MP6-RTE) are enriched with eggs, the main reason for this meal’s high cholesterol content. Interestingly, the ingredient containing cholesterol in the chicken drumstick (MP16-FF) and chicken wing (MP17-FF) samples is whey (10.3 mg cholesterol per cup).^52^ Additionally, for MP16-FF and MP17-FF, there’s a potential accumulation of these ingredients during the frying step depending on how frequently the frying oil of these fast food is replaced. In our previous study, we characterized the fatty acids composition of both RTE and FF classes ^36^. Herein, to help further discussion in oxidative status of UPFs, based on our previous data, we computed a series of fatty acids nutritional indexes as reviewed by Chen and Liu.^53^ (**Fig. 2A**). The PUFA/SFA index is used to determine the impact of fatty acids on cardiovascular health, since a diet rich in PUFA has been linked to lower serum cholesterol levels.^53^ In the present study, no significant difference in PUFA/SFA index was observed between FF and RTE. The indexes of atherogenicity (IA) and thrombogenicity were both proposed in the early ‘90s and are linked to specific nutritional attributes of fatty acids. These indexes measure the atherogenic/thrombogenic potential of fatty acids, as a ratio between common saturated fatty acids and total unsaturated fatty acids.^54^ We calculate these indices for the UPFs profiled in this study (see **Material and Methods**). RTE had a significantly higher IA and IT index value compared to FF (**Fig. 2A**); dairy foods were the class that most contributed to the high IA and IT values (**Fig. 1B,C**), for their load of SFA.^36^ Finally, the HH (Hypocholesterolemic/Hypercholesterolemic ratio), the HPI (Health-Promoting Index) and the UI (Unsaturated Index) are higher for foods with higher health benefits. In our database, FF scored higher than RTE for all the three indexes (**Fig. 2A**).

### 3.2. Oxidative status of the UPFs

The thiobarbituric acid reactive substances assay (TBARs) is widely used to measure oxidative stress in biological samples^55^ as well as foods.^56^ Lipid peroxidation results in the formation of conjugated dienes, lipid hydroperoxides and degradation products such as alkanes, aldehydes and isoprostanes. The formed hydroperoxides are further decomposed to flavorful secondary oxidation products, which are mainly aldehydes, such as hexanal, 4-hydroxynonenal (HNE), and malondialdehyde (MDA). The TBARs method detects lipid peroxidation products, such as malondialdehyde (MDA), a final product of fatty acids peroxidation. In this study, MDA concentration values were not significantly different between FF and RTE, but significant differences were found between UPFs’ categories (**Fig. 3A**). The highest values for significant amounts of MDA were observed in Meat and Poultry category, followed by Baby Food and Others category (**Table S1 Supplemental information**). Notability, in several samples, MDA was not detected. It could be possible that second stage oxidation molecules were not either present or the sample was at later stages of the oxidative chain reaction, in which complex and non-reactive species are formed and unable to be detected.

**Figure 3.**
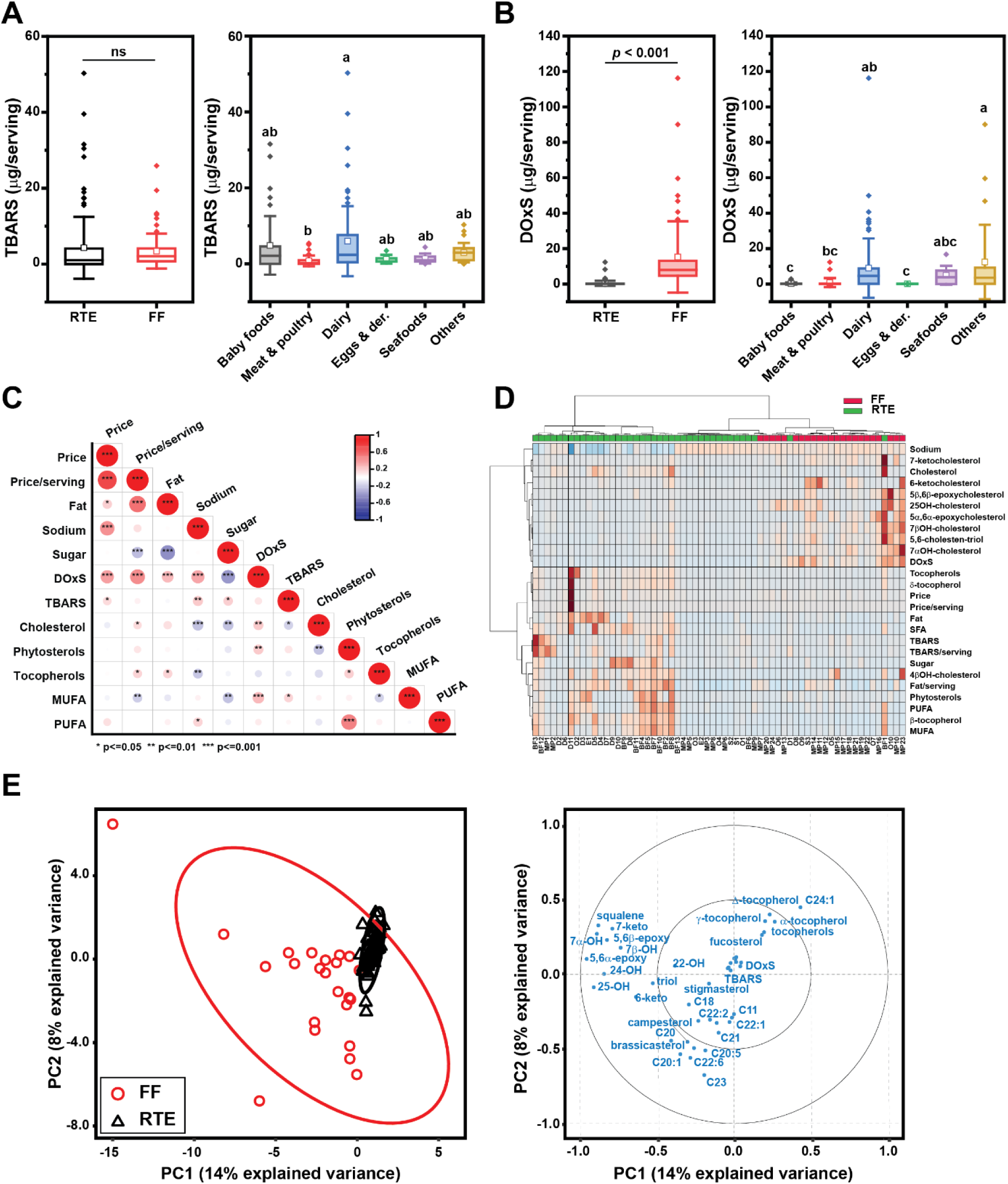
Oxidative status of UPFs contents in UPFs by group and category. (A) TBARs values; (B) DOxS amounts; (C) Pearson correlation between oxidative status and selected compositional variables; (D) heat-maps showing clustering of RTE and FF according to oxidative markers and selected classes of lipids; (E) principal components analysis of lipids and sterols, showing clustering of RTE and FF (left), as well as variables loadings (right).When comparing RTE and FF a paired *t*-test was used. The grouping letters in the graph represent distinct categories within the variable under analysis, highlighting significant differences between the groups as determined by the ANOVA, followed by post-hoc Tukey test (p<0.05).

We then quantified the total amount of DOxS, which reports cholesterol oxidation. DOxS are cholesteryl-derivatives that accumulate in significant amount within food matrices because of food manufacturing. Importantly, DOxS have known biological effects in both *in vitro* and animal models.^57, 58^ The FF group showed lower DOxS content (0.15-0.67 µg/serving 95% CI) compared to RTE (10.5-20.1 µg/serving 95% CI) (**Fig. 3B**). As expected, dairy foods contained higher amounts of DOxS because they are rich in cholesterol.^42, 59, 60^ DOxS content was positively correlated with price (both absolute and *per serving* base), as well as fat and sodium content (*p* < 0.001) (**Fig. 3C**). Conversely, the negative correlation of those macronutrients with cholesterol content reaffirms that DOxS accumulation is indeed driven by food processing.

### 3.3 DOxS quantification in UPFs

Twelve DOxS (7α-OH, 7β-OH, 4β-OH, 5,6α-epoxy, 5,6β-epoxy, triol, 6-keto, 7-keto, 20α-OH, 22-OH, 24-OH, and 25-OH) were identified in FF meals and RTE foods. The DOxS content for individual food samples is reported in the Supplemental Material (**Table S4-S5 Supplemental material**). DOxS content were significantly higher in the O group, followed by MP, and S (**Table 1**). 7α-OH was the most abundant DOxS in all UPFs, followed by 7β-OH. Notably, 4β-OH was only detected in FF meals. Also, two side-chain COPs (22-OH and 25-OH) were the most abundant specifically in baby food samples. The heat-map in **Fig. 3D** highlights compositional differences between FF and RTE, which were better resolved using sparse Partial-Least Square-Discriminant Analysis (sPLS-DA) (**Fig. 3E**). For the sPLS-DA we included the FAME data from our previous study published somewhere else ^36^. The multivariate analysis showed that RTE are tightly grouped when considering reduced variables featuring lipid and sterols composition. On the other hand, FF were broadly distributed across a dimension comprising several DOxS, but also long-chain FAME and phytosterols. These differences in oxidative load hints that specific compounds may serve as potential biomarkers for processing.

#### 3.2.1. Baby Food (BF)

Large quantities of phytosterols and cholesterol in BFs meals are intended to fulfill the infant’s nutritional needs during this critical period when their body - especially the brain - is under growth and development. For BF, the ingredients used during their confections such as meat and plant-based oils (i.e., soy, coconut, palm, or sunflower oils, etc.) could be beneficial for infants based on their phytosterols and cholesterol specific nutritional needs. However, infants’ diet should be closely monitored to avoid uncontrolled accumulation of these compounds that could result in serious health problem such as accelerated atherosclerosis – a consequence of an increased absorption of phytosterols and cholesterol in infants with sitosterolemia ^61^, or the development of parenteral nutrition-associated liver disease which is related to a higher intake of phytosterols ^62^.

In addition to phytosterols and cholesterol, we found 0.31 mg per serving of DOxS ingested by infants that is obtained through these same ingredients. Potentially, DOxS could negatively affect the infant’s health in the long run because of their accumulation through this specific diet. Low concentrations of DOxS (1 nM) have been reported in studies of different cardiometabolic, neurodegenerative, inflammatory bowel, ocular diseases, and have been even related to ageing regulation ^63–65^. Unfortunately, even though there’s abundant information regarding DOxS and adverse health effects on adults, little is known about their relationship with infants’ health and their diet.

Maldonado and coworkers performed a dietary exposure assessment of DOxS in BF and infant formulas (IF) after their consumption throughout the first year of life ^66^ (Maldonado et al, 2022). They confirmed the dominant presence of cholesterol and phytosterols amounts ingested and absorbed by this specific population group. 5,6β-epoxycholesterol, 7β-OH, and 7α-OH were the highest DOxS exposed through infants’ diet with 1,244.4, 191.8, and 165.5 µg/day, respectively; but 5,6β-epoxycholesterol was the most absorbed DOxS with a value of 0.040 mg/kg-day ^66^. Further studies focused on DOxS and their effect on infant’s health are required to address this knowledge gap.

#### 3.2.2. Eggs and Its Derivatives

For years, the recommended cholesterol daily intake was 300mg/day, and people took close attention to their egg’s daily consumption. Recently, this value was eliminated from the 2015-2020 Dietary Guidelines for Americans, the 2016 Japanese Dietary Guidelines, and the 2016 Chinese Dietary Guidelines ^67^ because of the lack of evidence of an overall positive association between egg consumption and risk of coronary heart disease ^68, 69, 70^. Even though, dietary cholesterol intake raises plasma cholesterol and LDL cholesterol, the effect is not as significant as originally thought, and its consumption also provides other nutrients such as proteins, unsaturated fats, folate, B vitamins, and minerals which are beneficial to human health (Hu, 2017).

Eggs and derivatives emerged as the second-highest category with respect to secondary oxidation products (MDA), indicating a higher extent of lipid oxidation in comparison to other categories. Initially, we hypothesized a positive correlation between MDA levels and DOxS (as observed in the Meat and Poultry category, which will be discussed in the next section). However, for the Eggs category, this was not the case, as it exhibited the lowest content of DOxS among all the groups.

#### 3.2.3. Meat and poultry (MP)

MP had the highest quantities of MDA compared to the other categories. These results were obtained mainly from samples made of pork and chicken. As we mentioned above, MP and E categories showed an opposite association related to DOxS versus MDA results. MP showed a higher DOxS content with high MDA values, while E showed the lowest DOxS content with high MDA values. As we mentioned before, this behavior could be caused by the presence of phytosterols in the sample - MP had significantly lower phytosterols’ contents-because of the addition of other ingredients to the whole meal. MDA values are aligned with previous studies ^71–75^ that report lipid oxidation changes in food samples after the addition of different ingredients and spices, with considerable amounts of phytochemicals that hinder the lipid oxidation process in the sample.

#### 3.2.4. Dairy products

Our initial hypothesis suggested that the lower presence of phytosterols in the Eggs category would lead to a higher DOxS content, as these phytosterols may have less antioxidant effect on cholesterol, resulting in the formation of more DOxS. However, our findings contradicted this hypothesis. Interestingly, the processing of dairy products involves subjecting them to very high temperatures, but the duration of these heating steps is considerably shorter (e.g., pasteurization: 17-20 seconds) compared to other cooking processes used for the samples in our study (e.g., frying: 10-15 minutes). Previous research has indicated that when comparing high cooking temperatures with short cooking times against low cooking temperatures with longer cooking times, there is typically less formation of different DOxS. Hashari and co-workers reported 360.5 ug 7β-OH/100 g sample of sausage for oven roasting (150℃) for 15min, but 204.3 ug 7β-OH/100 g sample sausage for microwave cooking (350℃) for 2min.^76, 77^ Moreover, Alina and co-workers detected 2.28 ppm of 5,6β-Epoxy after oven roasting mutton samples (190℃) for 25 min, but 0.83 ppm of 5,6β-Epoxy after grilling (230℃) for 20 min.^78^

Lastly, the D category was one of the only two categories where tocopherols could be detected in. It is possible that they can demote certain types of cancer, heart disease, and other chronic ailments.^79–81^ They are commonly found in vegetables and their oils, but traces of tocopherols were found in the D category of this study. Their presence is potentially associated with the use of vegetable colors and other preservatives used during dairy products manufacturing.

#### 3.2.5. Seafood

Seafood is known as one of the main sources of several essential lipids (ω-3 fatty acids, vitamins-D, and vitamin B12) which are crucial to different biological functions of the human body ^82^, and exert nutraceutical properties ^83^. In 2017, the Center of the Plate: Seafood and Vegetarian Consumer Trend Report states that 36% of adults are eating more seafood in place of meat.^84^ Even though the seafood category contains the lowest cholesterol and phytosterols amounts, it ranked on the 3^rd^ position of DOxS content (**Fig. 2**). Results are comparable with previous studies.^85–89^ The small number of foods used in this category may be the reason for the observed results.

#### 3.2.6. Other products (O)

The “Others” products category ranked 1st in both tocopherols and DOxS contents, as shown in **Fig. 2**. The higher presence of tocopherols in this category can be attributed to the significant use of vegetable oils during the preparation of these meals. Interestingly, while DOxS have been detected in other foods such as biscuits and French fries (especially those from European countries) according to a study in 2002,^90^ no research has reported their content in the same products available in the Western Diet. It is well-established that high cooking temperatures (ranging from 180°C to 200°C) and the specific type of oil used during deep frying of French fries are primary factors that promote the formation of DOxS. These findings highlight the importance of considering both the cooking methods and the choice of ingredients, such as oils, in the context of dietary habits, as they can significantly influence the formation of potentially harmful compounds like DOxS in our food.

### 3.4 DOxS as biomarkers of food processing: a preliminary assessment

We looked for specific lipid oxidative biomarkers for differentiating food belonging to the FF and RTE categories. To improve discriminatory power, we used a multivariate exploratory ROC analysis to assist the identification of a minimum number of features to use in the model. The model predicts high power discrimination between FF and RTE even when using only to features (AUC = 0.843), whereas the highest power is achieved with 10 features (AUC = 0.949) (**Fig. 4A**). The accuracy is >75% for all the considered models (**Fig. 4B**). From the VIP plot (**Fig. 4C**), a few DOxS were selected as top features, including 25-OH, triol and 7-keto, which were previously highlighted in the heatmap (**Fig. 3D**). Finally, using the first 2 features (25-OH and triol), we achieved sufficient discrimination between FF and RTE, being only 5 food items (three meat products, one baby food and a miscellaneous) assigned to the wrong category.

**Figure 4.**
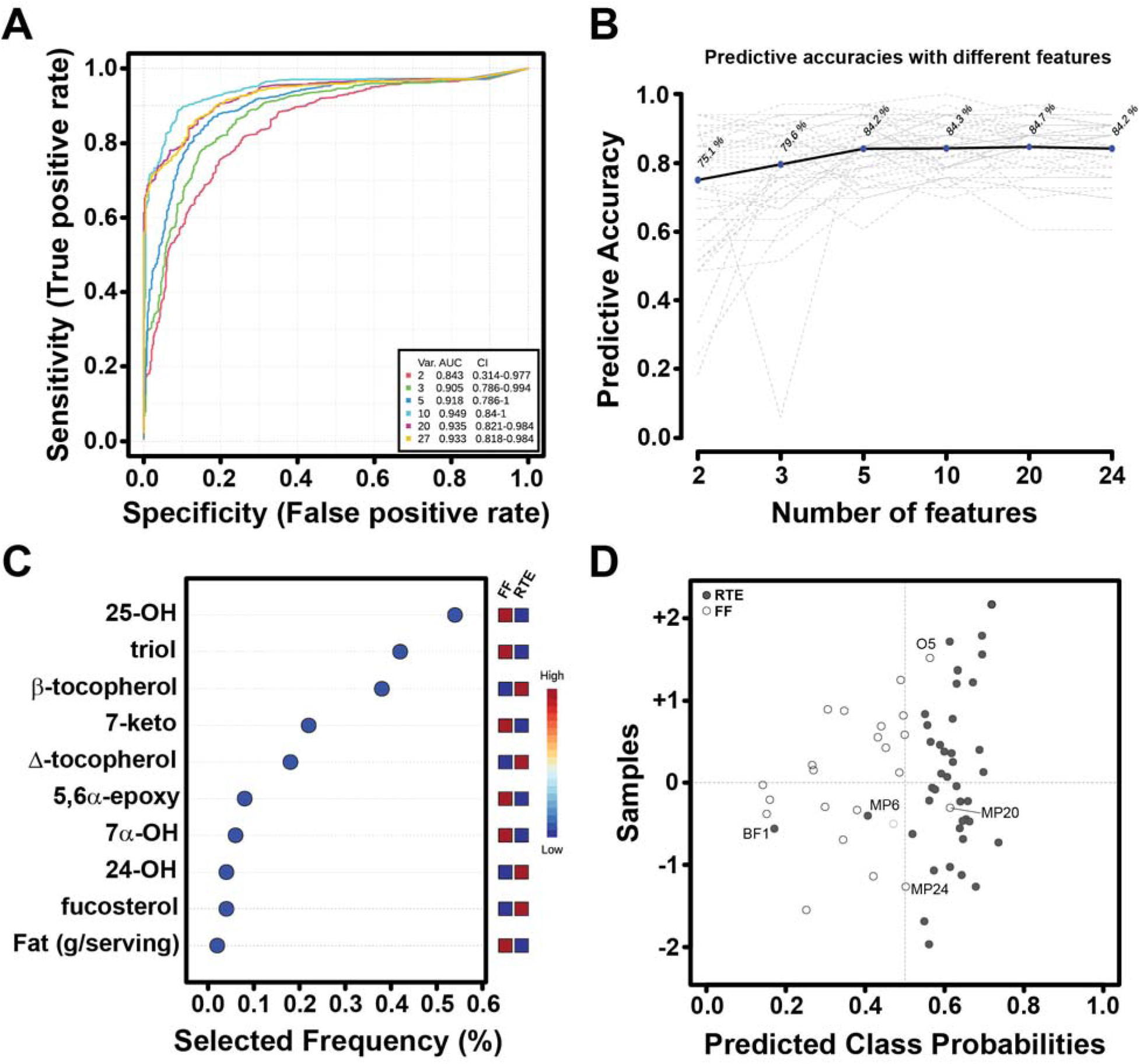
Biomarker analysis for UPFs. (A) ROC curves based on cross validation performance using a SVM classification method and a built-in SVM feature ranking method. (B) Predictive accuracies with increasing number of features. (C) Top ten significant features with selected frequencies for RTE and FF groups. (D) Predicted class probabilities as a result of average cross-validation for each sample using the best classifier (according to the AUC value). Mis-classified samples are labeled.

Overall, our results demonstrate clear differences in the oxidative status of FF and RTE, particularly when referring to DOxS.

## 4. Discussion

The use of the ambiguous term “ultra-processed foods” in the NOVA classification has given rise to misleading perceptions among consumers and the scientific community regarding the advantages of food preservation technologies. Numerous studies have linked the consumption of UPFs with adverse health outcomes, but there is a lack of quantitative data or biomarkers to assess the “level” of processing in these food items. Moreover, the variability in manufacturing processes and the sample matrix further complicate the analysis of food composition and its metabolites. In this study, we present an initial evaluation of the oxidative status of UPFs using an onsite database and quantitative lipidomics focusing on the main lipid oxidative species.

### FF and RTE have distinct oxidative signatures

We previously reported that RTE meals not only have higher fat content compared to FF meals, but also a different FAME composition.^36^ These differences are due to the extensive formulation and ingredients used to produce more palatable and tasty meals in addition to the cooking and preservation methods employed during their manufacturing. In the present study, we confirmed that formulation also affects the sterols composition of UPFs (**Fig. 1**), and most importantly their oxidative status. Although both groups (FF and RTE) are considered UPFs, there is a clear distinction between these meals due to type of transformation,^91^ ^enzymatical^ source of lipid and protein (animal-based vs plant-based), type of preservation technology (e.g., thermal vs nonthermal, wet versus dry), additives addition (e.g, edulcorates, colorants, acidifies). The question that raises is if the differences in composition and manipulation, from both a manufacturing and preparation point of view, will be reflected in major lipid oxidative markers. For instance, although no difference was observed in secondary oxidative products (measured as MDA), FF accumulates more DOxS, which are derived from chemical oxidation of cholesterol (**Fig. 2A,B**).

### DOxS, UPFs, Nutrition and Public Health

The recent alterations to dietary guidelines worldwide stem from emerging research findings. Some of these studies propose that cholesterol alone may not be the sole culprit responsible for chronic diseases like CHD. Instead, they indicate that various other factors, such as overall dietary patterns and their impact on the gut microbiota, play a significant role in the development of human pathological conditions. Recent studies have reported association of specific gut metagenomic species with multiple plasma metabolites.^92–99^ Characterization of these interactions was stored in an online atlas created by Dekkers and coworkers,^100^ expecting to help the scientific community in the process of understanding the effects of the gut microbiota on health It is possible that DOxS could be also influencing changes on the gut microbiota which could end up affecting human health.^101^

### Identification of oxidized biomarkers

**Table S6** (**Supplemental Information**) shows results of the biomarker analysis using MetaboAnalyst package.^102^ Brassicasterol was the potential biomarker for RTE meals, while 7α-OH and 7β-OH were both suggested for FF meals. While both groups fall under the category of UPFs, the distinct production and processing characteristics of each group have a notable influence on the biomarker analysis outcomes. Our findings suggest that brassicasterol and 7α-OH + 7β-OH may potentially serve as biomarkers for ready-to-eat (RTE) and fast-food (FF) meals, respectively. However, we acknowledge the significant impact of the diverse food categories on these results, and further investigations are required to validate these potential biomarkers. **Table S6** also shows potential biomarkers for each individual food category.

Maldonado and co-workers^66^ reported 7α-OH as a potential biomarker of the BF category after comparing it against powered and liquid IF (PIF and LIF, respectively) from Kilvington’s study.^26^ In our study, when comparing BF against all other categories, the biomarker analysis pointed to β-sitosterol as a potential biomarker for this specific category. However, we believe that 7α-OH could potentially be a more accurate biomarker, as it reflects the processing differences between these two types of infant foods. On the other hand, the presence of β-sitosterol in these meals may be more closely associated with the use of vegetable oils, which are key ingredients in many of these products. Thus, further investigation is needed to ascertain the most suitable biomarker for distinguishing between these food categories accurately. Further research quantifying DOxS in a broader dataset of UPFs found in the WD is suggested to increase the diet’s variability and evaluate any possible changes in the results obtained from the biomarker analysis. This expanded analysis will provide valuable insights into the specific biomarkers that can best differentiate between different categories of UPFs, contributing to a better understanding of their impact on human health.

Consumer awareness of foods and their healthiness is of critical importance,^103, 104^ and varies across culture. A pilot study performed in Europe found that Italian and Dutch consumers have weaker negative opinion towards UPF compared to Brazilian consumers.^103^ However, healthiness perception of UPF seems to be linked to heuristic assessment.^104^ For instance, the National Health Institute (NIH) recognizes the need for more data to provide insight into personalized foods with the program precision nutrition.^105^ A dietary assessment will enable the development of an exposure assessment that would become the starting point of a most-needed risk assessment to elucidate the association between these food meals and disease risk.^106^ Several dietary markers of exposure for many foods have been putatively identified.^107–110^

## 5. Conclusion

Our study revealed significant differences in DOxS content between RTE and FF meals. Similarly, the levels of phytosterols exhibited a significant difference between RTE and FF meals. These findings underscore the substantial influence of the parameters and conditions used in the manufacturing processes of these two groups of UPFs. Furthermore, the composition of the food matrix played a pivotal role in determining the presence of these bioactive lipids, as observed when comparing all six food categories. Overall, not all UPFs are deemed to be “unhealthy”, a lot of these compounds such as phytosterols and tocopherols are needed to avoid oxidative species that promote development or worsening of human illnesses. However, for RTE meals, there’s definitely room for improvement in finding a solution to avoid the excessesive addition of certain nutritional attributes such as sodium, sugar, and fat as part of the current “solutions” for different nutritional and/or safety processes (i.e., microbial reduction, preservation, shel-life extension, etc.). Meanwhile, FF meals seem to be a big source of DOxS. The intake of these toxicological compounds, raises even more health concerns from a chemical and molecular point of view regarding UPFs. Unfortunately, the presence of DOxS in UPFs increse the risk of developmeing of cardiometabolic disorders (CMD), which are one of the most challenging global health issues of the 21st century, recognized by the World Health Organization (WHO),^111^ and the American Society of Endocrinology.^112^ Oxidative compounds like these cholesterol oxidation products^113^, need to be avoided for the sake of the society’s health.

Therefore, we believe that the use of DOxS as biomarkers could potentially help in a future to identify in an early stage the presence of different chronic diseases, and even, prevent their development. Nevertheless, an exposure assessment is critical to understand the exposure level of these toxicological compounds – DOxS – and their relationship with CMD and other chronic diseases.

Until now, no nutritional database of UPFs has been fully developed yet. Maldonado and coworkers already analyzed the nutritional aspects of these same 63 UPFs evaluated in this study.^114^ Our results aim to expand that database which will serve as our contribution to the scientific community, and general society, in the ongoing quest of elucidating the relationship of DOxS and human health. The creation of a DOxS database of UPFs in WD developed in this study is just the first step to assess and determine the potential health risk caused by the consumption of these UPFs.

## Data Availability

All data produced in the present study are available upon reasonable request to the authors

## Acknowledgments

This study was partially funded by the Centre for Research Ingredients Safety (CRIS) of Michigan State University with the GR100229 grant, and the USDA National Institute of Food and Agriculture, Hatch project MICL02526 to I.G.M.M.

## Conflict of interest

Authors declare non conflict of interest for this study.

## Notes

### Competing Interest Statement

The authors have declared no competing interest.

