## Supplemental Table 1 for "Oxidative Status of the Ultra-Processed Foods in the Western Diet"

**Table S1:** MDA concentration in RTE items and FF meals.

| **Overall UPFs** | | | |
| --- | --- | --- | --- |
| ***Food Category*** | ***µg MDA/g fat ± STD*** | ***Kruskal-Wallis Test*** | |
| Dairy | 0.785 ± 1.42 | *p=0.0114* | |
| Meat & Poultry | 5.97 ± 9.23 |  |  |
| Baby Food | 4.86 ± 7.70 |  |  |
| Eggs and Derivatives | 1.20 ± 1.16 |  |  |
| Seafood | 1.40 ± 1.34 |  |  |
| Others | 2.95 ± 2.54 |  |  |
| **READY-TO-EAT** | | | |
| ***Food Category*** | ***Food Item*** | | ***µg MDA/g fat ± STD*** |
| Dairy | D1-RTE | | 3.58 ± 1.28 |
|  | D2-RTE | | 0.36 ± 0.23 |
|  | D3-RTE | | - |
|  | D4-RTE | | - |
|  | D5-RTE | | 1.03 ± 0.052 |
|  | D6-RTE | | 0.19 ± 0.017 |
|  | D7-RTE | | 0.37 ± 0.27 |
|  | D8-RTE | | - |
|  | D9-RTE | | - |
|  | D10-RTE | | 3.22 ± 2.18 |
|  | D11-RTE | | - |
|  | ***Average ± STD*** | | ***0.785 ± 1.39*** |
| ***Meat & Poultry*** | MP1-RTE | | 29.66 ± 8.43 |
|  | MP2-RTE | | 12.67 ± 3.42 |
|  | MP3-RTE | | 7.59 ± 0.23 |
|  | MP4-RTE | | 7.73 ± 5.87 |
|  | MP5-RTE | | 2.12 ± 1.48 |
|  | MP6-RTE | | - |
|  | MP7-RTE | | - |
|  | MP8-RTE | | 6.76 ± 4.83 |
|  | MP9-RTE | | 23.83 ± 18.69 |
|  | ***Average ± STD*** | | ***9.68 ± 12.48*** |
| ***Baby food*** | BF1-RTE | | - |
|  | BF2-RTE | | - |
|  | BF3-RTE | | 19.42 ± 7.41 |
|  | BF4-RTE | | - |
|  | BF5-RTE | | 3.30 ± 0.87 |
|  | BF6-RTE | | 7.95 ± 5.47 |
|  | BF7-RTE | | 2.47 ± 0.42 |
|  | BF8-RTE | | 2.81 ± 1.43 |
|  | BF9-RTE | | 1.48 ± 0.24 |
|  | BF10-RTE | | 9.36 ± 0.51 |
|  | BF11-RTE | | 2.51 ± 1.09 |
|  | BF12-RTE | | 17.87 ± 11.01 |
|  | BF13-RTE | | - |
|  | ***Average ± STD*** | | ***4.86*** ***± 7.61*** |
| Eggs and Derivatives | E1-RTE | | 0.56 ± 0.042 |
|  | E2-RTE | | 1.85 ± 1.19 |
|  | ***Average ± STD*** | | ***1.20 ± 1.06*** |
| Seafood | S1-RTE | | 1.26 ± 0.66 |
|  | ***Average ± STD*** | | ***0.84* *± 0.80*** |
| Others | O1-RTE | | 1.88 ± 1.46 |
|  | O2-RTE | | 3.03 ± 0.89 |
|  | O3-RTE | | 3.83 ± 0.60 |
|  | O4-RTE | | 1.24 ± 0.23 |
|  | ***Average ± STD*** | | ***2.49*** ***± 1.36*** |
| **Overall RTE Average ± STD** | | | ***4.30 ± 8.14*** |
| **FAST FOOD** | | | |
| ***Food Category*** | ***Food Item*** | | ***µg MDA/g fat ± STD*** |
| Meat & Poultry | MP10-FF | | 4.15 ± 1.14 |
|  | MP11-FF | | 1.35 ± 0.79 |
|  | MP12-FF | | - |
|  | MP13-FF | | 3.43 ± 1.70 |
|  | MP14-FF | | - |
|  | MP15-FF | | 3.64 ± 2.24 |
|  | MP16-FF | | - |
|  | MP17-FF | | - |
|  | MP18-FF | | 17.01 ± 6.32 |
|  | MP19-FF | | 2.13 ± 0.47 |
|  | MP20-FF | | 1.20 ± 0.71 |
|  | MP21-FF | | 4.36 ± 1.24 |
|  | MP22-FF | | 3.41 ± 1.95 |
|  | MP23-FF | | 1.53 ± 0.69 |
|  | MP24-FF | | 11.20 ± 7.77 |
|  | **Average ± STD** | | **3.75 ± 5.25** |
| Seafood | S2-FF | | 2.66 ± 1.25 |
|  | S3-FF | | 0.69 ± 0.38 |
|  | ***Average ± STD*** | | ***1.67 ± 1.35*** |
| Others | O5-FF | | 7.27 ± 2.61 |
|  | O6-FF | | 1.84 ± 1.64 |
|  | O7-FF | | 2.11 ± 0.90 |
|  | O8-FF | | - |
|  | O9-FF | | 3.22 ± 1.86 |
|  | O10-FF | | 6.20 ± 2.40 |
|  | **Average ± STD** | | **3.25** **± 2.99** |
| **Overall FF Average ± STD** | | | **3.44 ± 4.60** |

- Means No detected
