## Supplemental Table 2 for "Oxidative Status of the Ultra-Processed Foods in the Western Diet"

**Table S2:** Cholesterol content in UPFs by group and food category. (Total values per category are expressed in mean and range).

| ***READY-TO-EAT*** | | | | |
| --- | --- | --- | --- | --- |
| ***Food Category*** | ***Sample ID*** | ***Cholesterol***  ***(mg/100 g fat) ± STD*** | ***Cholesterol Reference***  ***(mg/100 g fat)*** | ***Serving Size*** |
| Dairy | D1-RTE | 20.30 ± 15.85 | 333.33 | 19.00 g |
|  | D2-RTE | 196.33 ± 29.43 | 333.33 | 21.00 g |
|  | D3-RTE | ND | N/A | 1 tbsp |
|  | *D4-RTE | 305.90 ± 41.16 | 272.73 | 14.00 g |
|  | D5-RTE | 165.09 ± 30.33 | 285.71 | 30.00 mL |
|  | D6-RTE | 287.08 ± 20.03 | 300.00 | 28 .00g |
|  | D7-RTE | 238.86 ± 133.50 | 285.71 | 28.00 g |
|  | D8-RTE | 243.02 ± 40.22 | 357.14 | 0.50 cup (65.00 g) |
|  | D9-RTE | 368.25 ± 72.77 | 0.00 | 1 container (99.00 g) |
|  | D10-RTE | 282.18 ± 52.16 | 500.00 | 414.03 g |
|  | D11-RTE | 16.25 ± 3.59 | NR | 1 scoop (9 g) |
|  | ***Total*** | **20.3 (0 – 368.25)** |  | |
| Baby food | BF1-RTE | 1,247.23 ± 227.45 | 1,000.00 | 71.00 g |
|  | BF2-RTE | 673.84 ± 98.50 | 833.33 | 4.00 oz (113.40 g) |
|  | BF3-RTE | 58.17 ± 8.50 | 250.00 | 4.00 oz (113.40 g) |
|  | BF4-RTE | 319.43 ± 83.74 | 500.00 | 4.00 oz (113.40 g) |
|  | BF5-RTE | 348.65 ± 82.26 | 400.00 | 4.00 oz (113.40 g) |
|  | BF6-RTE | 123.24 ± 29.61 | 7.81 | 128.00 g |
|  | BF7-RTE | 263.77 ± 76.69 | 400.00 | 4.00 oz (113.40 g) |
|  | BF8-RTE | 683.55 ± 310.63 | 875.00 | 71.00 g |
|  | BF9-RTE | 162.73 ± 95.67 | 333.33 | 4.00 oz (113.40 g) |
|  | BF10-RTE | 251.02 ± 57.95 | 333.33 | 4.00 oz (113.40 g) |
|  | BF11-RTE | 313.34 ± 27.54 | 285.71 | 4.00 oz (113.40 g) |
|  | BF12-RTE | 172.87 ± 17.84 | 333.33 | 4.00 oz (113.40 g) |
|  | BF13-RTE | 327.40 ± 87.35 | 500.00 | 85.00 g |
|  | ***Total*** | **380.40 (50.17 – 1,247.23)** |  | |
| Meat & Poultry | MP1-RTE | 224.75 ± 15.73 | 333.33 | 38.00 g |
|  | MP2-RTE | 648.56 ± 125.37 | 250.00 | 32.00 g |
|  | MP3-RTE | 327.69 ± 28.27 | 200.00 | 240.00 mL |
|  | MP4-RTE | 157.80 ± 55.79 | 500.00 | 1 can (236.59 g) |
|  | MP5-RTE | 396.25 ± 46.38 | 500.00 | 11.00 oz (227.00 g) |
|  | MP6-RTE | 2,023.90 ± 209.36 | 750.00 | 0.5 cup (113.40 g) |
|  | MP7-RTE | 352.50 ± 166.24 | 583.33 | 0.5 cup (240.00 g) |
|  | MP8-RTE | 62.55 ± 8.88 | 142.86 | 1 cup (249.00 g) |
|  | MP9-TE | 199.16 ± 96.55 | 181.82 | 1 cup (257.00 g) |
|  | ***Total*** | **488.13 (62.55 – 2023.90)** |  | |
| Seafood | S1-RTE | 43.89 ± 1.81 | 50.00 | 18.80 oz (532.97 g) |
|  | ***Total*** | **43.89** |  | |
| Eggs and derivatives | E1-RTE | 98.44 ± 16.35 | 90.91 | 1 tbsp |
|  | E2-RTE | 274.11 ± 40.94 | 218.75 | 0.50 cup (115.00 g) |
|  | ***Total*** | **186.28 (98.44 – 274.11)** |  | |
| Others | O1-RTE | ND | NR | 2 tbsp (30.00 g) |
|  | O2-RTE | 51.55 ± 11.01 | 66.67 | 2 tbsp (30.00g) |
|  | O3-RTE | 97.51 ± 32.67 | 333.33 | 2.5 oz (70.90 g) |
|  | O4-RTE | 37.95 ± 8.46 | 285.71 | 1 package (58.00 g) |
|  | ***Total*** | **62.34 (37.95 – 97.51)** |  | |
| ***FAST FOOD*** | | | | |
| ***Food Category*** | ***Sample ID*** | ***Cholesterol***  ***(mg/100 g fat) ± STD*** | ***Cholesterol Reference***  ***(mg/100 g fat)*** | ***Serving Size*** |
| Meat & Poultry | MP10-FF | 370.80 ± 55.74 | 267.22 | 95.00 g |
|  | MP11-FF | 229.99 ± 53.08 | 222.05 | 4 pieces (64.00 g) |
|  | MP12-FF | 362.03 ± 8.90 | 296.86 | 119.00 g |
|  | MP13-FF | 446.38 ± 73.73 | 464.29 | 1 taco (102.00 g) |
|  | MP14-FF | 299.73 ± 46.37 | 230.77 | 1 quesadilla (170.00 g) |
|  | MP15-FF | 504.08 ± 7.82 | 245.31 | 1 burrito (140.00 g) |
|  | MP16-FF | 742.86 ± 62.13 | 820.56 | 1 piece (75.00 g) |
|  | MP17-FF | 728.22 ± 42.86 | 600.00 | 1 piece (60.00 g) |
|  | MP18-FF | 333.59 ± 105.66 | 171.43 | 5.40 oz (153. 09 g) |
|  | MP19-FF | 310.55 ± 147.36 | 1,423.08 | 5.70 oz (161.69 g) |
|  | MP20-FF | 526.02 ± 61.70 | 312.92 | 1 sandwich (187.00 g) |
|  | MP21-FF | 450.17 ± 13.91 | 272.73 | 1 sandwich (71.28 g) |
|  | MP22-FF | 335.74 ± 109.38 | 240.74 | 1 sandwich (94.61 g) |
|  | MP23-FF | 280.77 ± 38.10 | NA | 1 slice (123.00 g) |
|  | MP24-FF | 266.45 ± 30.50 | 200.00 | 1 slice (79.00 g) |
|  | ***Total*** | **387.21 (8.00 – 742.86)** |  | |
| Seafood | S2-FF | 216.98 ± 26.53 | 204.39 | 1 sandwich (131.00 g) |
|  | S3-FF | 405.14 ± 37.49 | 555.56 | 5.00 oz (141.75 g) |
|  | ***Total*** | **311.06 (216.98 – 405.14)** |  | |
| Others | O5-FF | ND | 0.00 | 1 medium serving (117.00 g) |
|  | O6-FF | ND | 222.99 | 1 biscuit (76.00 g) |
|  | O7-FF | 268.72 ± 43.02 | 0.00 | 3 hotcakes (149.00 g) |
|  | O8-FF | 16.42 ± 8.31 | 5.87 | 1 biscuit (49.00 g) |
|  | O9-FF | ND | 0.00 | 1 order (34.99 g) |
|  | O10-FF | 90.13 ± 21.47 | 0.00 | 1 order (16.85 g) |
|  | ***Total*** | **62.55 (16.42 – 268.72)** |  | |

ND = not detected

N/A = not apply

NR = not reported
