## Supplemental Table 3 for "Oxidative Status of the Ultra-Processed Foods in the Western Diet"

**Table S3:** Phytosterol content in UPFs. (Total values of phytosterols are expressed as mean and range.)

| ***Ready to Eat*** | | | | | | | |
| --- | --- | --- | --- | --- | --- | --- | --- |
| ***Food Category*** | ***Food Item*** | ***Brassicasterol*** | ***Campesterol*** | ***Stigmasterol*** | | ***β-Sitosterol*** | ***Fucosterol*** |
|  |  |  | | | ***(mg/100 g fat) ± STD*** | | |
| *Dairy* | D1-RTE | ND | 26.01 ± 9.31 | 20.30 ± 6.01 | | 86.64 ± 12.43 | ND |
|  | D2-RTE | ND | ND | ND | | ND | ND |
|  | D3-RTE | ND | 33.31 ± 14.70 | 43.37 ± 12.51 | | 136.05 ± 5.72 | ND |
|  | D4-RTE | ND | ND | ND | | ND | ND |
|  | D5-RTE | ND | ND | ND | | ND | ND |
|  | D6-RTE | ND | ND | ND | | ND | ND |
|  | D7-RTE | ND | ND | ND | | ND | ND |
|  | D8-RTE | ND | ND | ND | | ND | ND |
|  | D9-RTE | ND | ND | ND | | ND | ND |
|  | D10-RTE | ND | ND | ND | | ND | ND |
|  | D11-RTE | ND | 18.19 ± 4.86 | 11.64 ± 5.32 | | 95.02 ± 24.75 | ND |
|  | **Total** | **-** | **26.01 (18.19 – 33.31)** | **25.10 (11.64 – 43.37)** | | **105.90 (86.64 – 136.05)** | **-** |
| *Meat & Poultry* | MP1-RTE | ND | ND | ND | | ND | ND |
|  | MP2-RTE | ND | ND | ND | | tr | ND |
|  | MP3-RTE | ND | 68.45 ± 13.99 | 259.84 ± 40.55 | | 626.48 ± 57.01 | 58.18 ± 26.67 |
|  | MP4-RTE | ND | ND | 31.84 ± 9.45 | | 102.34 ± 29.25 | ND |
|  | MP5-RTE | ND | 14.60 ± 5.20 | ND | | ND | 0.033 ± 0.026 |
|  | MP6-RTE | ND | ND | ND | | ND | ND |
|  | MP7-RTE | ND | ND | ND | | ND | ND |
|  | MP8-RTE | ND | 36.47 ± 24.74 | tr | | 83.75 ± 43.50 | ND |
|  | MP9-RTE | ND | ND | ND | | ND | ND |
|  | **Total** | **-** | **39.84 (14.60 – 68.45)** | **145.84 (31.84 – 259.84)** | | **270.86 (83.75 – 626.48)** | **29.11 (0.03 – 58.18)** |
| *Seafood* | S1-RTE | ND | 33.99 ± 26.75 | tr | | 51.87 ± 37.21 | ND |
|  | **Total** | **-** | **33.99** | **tr** | | **51.87** | **-** |

**Table S3:** Phytosterol content in UPFs (cnt’d). (Total values of phytosterols are expressed as mean and range.)

| ***Ready to Eat*** | | | | | | |
| --- | --- | --- | --- | --- | --- | --- |
| ***Food Category*** | ***Food Item*** | ***Brassicasterol*** | ***Campesterol*** | ***Stigmasterol*** | ***β-Sitosterol*** | ***Fucosterol*** |
|  |  |  | ***(mg/100 g fat) ± STD*** | | | |
| *Eggs & egg’s derivatives* | E1-RTE | ND | 61.64 ± 24.67 | 42.06 ± 13.92 | 176.90 ± 18.66 | ND |
|  | E2-RTE | ND | 53.46 ± 6.89 | 37.17 ± 9.46 | 162.59 ± 24.23 | ND |
|  | **Total** | **-** | **57.55 (53.46 – 61.64)** | **39.62 (37.17 – 42.06)** | **169.75 (162.59 – 176.90)** | **-** |
| *Baby foods* | BF1-RTE | ND | ND | ND | ND | ND |
|  | BF2-RTE | ND | ND | ND | ND | ND |
|  | BF3-RTE | ND | 10.48 ± 5.14 | 6.52 ± 1.87 | 47.10 ± 13.21 | ND |
|  | BF4-RTE | ND | 182.76 ± 53.20 | 75.16 ± 41.86 | 498.98 ± 129.71 | ND |
|  | BF5-RTE | ND | 136.48 ± 47.29 | 33.77 ± 7.19 | 391.77 ± 93.78 | ND |
|  | BF6-RTE | ND | 21.98 ± 6.84 | ND | 100.31 ± 19.76 | ND |
|  | BF7-RTE | ND | 190.88 ± 21.15 | ND | 421.59 ± 137.27 | ND |
|  | BF8-RTE | ND | ND | ND | ND | ND |
|  | BF9-RTE | ND | ND | ND | ND | ND |
|  | BF10-RTE | ND | 48.06 ± 9.69 | 25.41 ± 14.33 | 160.14 ± 26.56 | ND |
|  | BF11-RTE | 19.72 ± 3.21 | ND | 48.31 ± 2.23 | 435.62 ± 41.06 | ND |
|  | BF12-RTE | 29.60 ± 3.30 | 99.33 ± 24.73 | ND | 236.18 ± 18.19 | ND |
|  | BF13-RTE | ND | 44.86 ± 3.55 | ND | ND | 0.046 ± 0.014 |
|  | **Total** | **24.66 (19.72 – 29.60)** | **91.85 (10.48 – 190.88)** | **37.83 (6.52 – 75.16)** | **286.46 (47.10 – 498.98)** | **0.046** |
| *Others* | O1-RTE | ND | 34.54 ± 5.32 | ND | ND | ND |
|  | O2-RTE | ND | 50.87 ± 14.28 | 38.09 ± 10.27 | 148.17 ± 8.02 | ND |
|  | O3-RTE | ND | 38.65 ± 19.12 | ND | ND | 5.45 ± 1.58 |
|  | O4-RTE | ND | 26.01 ± 9.31 | 16.34 ± 9.22 | 124.15 ± 12.47 | 39.85 ± 17.26 |
|  | **Total** | **-** | **37.52 (26.01 – 50.87)** | **27.22 (16.34 – 38.09)** | **136.16 (124.15 – 148.17)** | **22.65 (5.45 – 39.85)** |

**Table S3:** Phytosterol content in UPFs (cnt’d). (Total values of phytosterols are expressed as mean and range.)

| ***Fast Food*** | | | | | | |
| --- | --- | --- | --- | --- | --- | --- |
| ***Food Category*** | ***Food Item*** | ***Brassicasterol*** | ***Campesterol*** | ***Stigmasterol*** | ***β-Sitosterol*** | ***Fucosterol*** |
|  |  |  | ***(mg/100 g fat) ± STD*** | | | |
| *Meat & Poultry* | MP10-FF | ND | ND | ND | ND | ND |
|  | MP11-FF | ND | 122.85 ± 18.26 | ND | 296.67 ± 38.94 | ND |
|  | MP12-FF | ND | ND | ND | ND | ND |
|  | MP13-FF | ND | 66.46 ± 14.08 | 26.36 ± 10.01 | 176.24 ± 26.14 | ND |
|  | MP14-FF | ND | 81.36 ± 13.67 | 209.33 ± 40.88 | ND | ND |
|  | MP15-FF | ND | ND | 53.03 ± 7.46 | ND | ND |
|  | MP16-FF | ND | 86.20 ± 33.23 | ND | 165.55 ± 63.66 | ND |
|  | MP17-FF | 27.56 ± 4.66 | 101.79 ± 8.40 | ND | 173.08 ± 17.49 | ND |
|  | MP18-FF | ND | 68.98 ± 23.39 | 48.25 ± 13.69 | 343.35 ± 137.26 | ND |
|  | MP19-FF | ND | 4.77 ± 3.44 | ND | 33.40 ± 17.52 | ND |
|  | MP20-FF | ND | 22.44 ± 7.49 | 8.93 ± 7.17 | 95.24 ± 9.40 | ND |
|  | MP21-FF | ND | ND | ND | 39.39 ± 6.79 | ND |
|  | MP22-FF | ND | 20.69 ± 10.72 | ND | 34.28 ± 4.46 | ND |
|  | MP23-FF | ND | 83.95 ± 7.74 | 168.10 ± 24.33 | ND | ND |
|  | MP24-FF | ND | 23.57 ± 7.87 | ND | 68.94 ± 6.79 | ND |
|  | **Total** | **27.56** | **62.10 (4.77 – 122.85)** | **85.67 ( 8.93 – 209.33)** | **142.61 (33.40 – 343.35)** | **-** |
| *Seafood* | S2-FF | ND | 60.10 ± 22.86 | ND | 199.39 ± 26.35 | ND |
|  | S3-FF | ND | 25.42 ± 4.88 | 19.73 ± 13.05 | 36.94 ± 4.58 | ND |
|  | **Total** | **-** | **42.76 ( 25.42 – 60.10)** | **19.73** | **118.17 (36.94 – 199.39)** | **-** |
| *Others* | O5-FF | ND | 192.00 ± 21.55 | ND | 331.87 ± 34.11 | ND |
|  | O6-FF | ND | ND | 36.45 ± 17.21 | ND | ND |
|  | O7-FF | ND | 40.86 ± 5.16 | 224.87 ± 30.79 | ND | ND |
|  | O8-FF | ND | 35.93 ± 23.68 | ND | 74.36 ± 8.20 | ND |
|  | O9-FF | 44.19 ± 17.67 | 184.08 ± 25.17 | ND | 333.41 ± 45.98 | ND |
|  | O10-FF | ND | 60.98 ± 12.72 | ND | 102.64 ± 19.30 | tr |
|  | **Total** | **44.19** | **102.77 (35.93 – 192.00)** | **130.66 (36.45 – 224.87)** | **210.57 (74.36 – 333.41)** | **tr** |

ND = not detected, tr = traces
