## Supplemental Table 4 for "Oxidative Status of the Ultra-Processed Foods in the Western Diet"

**Supplemental material**

**Table S4:** DOxS content in RTE group, expressed in mg per 100g of fat.

|  | ***Dairy*** | | | | | | | | | | | | | |
| --- | --- | --- | --- | --- | --- | --- | --- | --- | --- | --- | --- | --- | --- | --- |
|  | | ***7α-OH*** | ***7β-OH*** | ***4β-OH*** | ***5,6α-Epoxy*** | ***5,6β-Epoxy*** | ***7-Keto*** | ***Triol*** | ***6-Keto*** | ***20α-OH*** | ***22-OH*** | ***24-OH*** | ***25-OH*** | ***Total COPs*** |
|  | ***(mg/100 g fat) ± STD*** | | | | | | | | | | | | | |
| ***D1-RTE*** | | 0.15 ± 0.049 | 0.14 ± 0.080 | - | - | - | - | 0.0 | - | - | - | - | 1.05 ± 1.21 | 63.59 ± 25.32 |
| ***D2-RTE*** | | 0.084 ± 0.034 | 0.086 ± 0.026 | - | - | - | 0.044 ± 0.029 | 0.051 ± 0.0050 | - | - | - | - | - | 0.55 ± 0.13 |
| **D3-RTE** | | - | - | - | - | - | - | - | - | - | - | - | 0.053 ± 0.020 | 0.14 ± 0.048 |
| **D4-RTE** | | 0.64 ± 0.29 | 0.51 ± 0.21 | - | 0.044 ± 0.025 | 0.050 ± 0.019 | 0.37 ± 0.13 | 0.18 ± 0.12 | 0.046 ± 0.036 | - | - | - | - | 2.00 ± 0.92 |
| **D5-RTE** | | 0.19 ± 0.0061 | 0.12 ± 0.029 | - | - | - | 0.11 ± 0.016 | - | - | - | - | - | - | 1.29 ± 0.39 |
| **D6-RTE** | | 0.32 ± 0.10 | 0.27 ± 0.036 | - | - | - | 0.11 ± 0.025 | 0.051 ± 0.0056 | - | - | - | - | - | 1.72 ± 0.26 |
| **D7-RTE** | | 0.51 ± 0.26 | 0.39 ± 0.20 | - | 0.023 ± 0.0046 | 0.058 ± 0.011 | 0.20 ± 0.094 | 0.14 ± 0.032 | 0.046 ± 0.020 | - | - | - | - | 1.41 ± 0.52 |
| **D8-RTE** | | 0.039 ± 0.019 | 0.043 ± 0.018 | - | - | - | - | - | - | - | - | - | - | 0.21 ± 0.081 |
| **D9-RTE** | | 0.36 ± 0.056 | 0.35 ± 0.072 | - | - | - | 0.16 ± 0.020 | 0.10 ± 0.013 | 0.065 ± 0.038 | - | - | - | - | 4.06 ± 0.98 |
| ***D10-RTE*** | | 2.75 ± 1.88 | 4.72 ± 3.26 | - | 0.20 ± 0.16 | - | 1.40 ± 0.92 | 2.49 ± 1.84 | 0.19 ± 0.038 | - | 0.15 ± 0.068 | - | 0.43 ± 0.26 | 16.09 ± 8.01 |
| ***D11-RTE*** | | 0.075 ± 0.0094 | 0.09 ± 0.033 | - | - | - | - | - | - | 0.055 ± 0.032 | - | - | - | 0.90 ± 0.061 |

|  | ***Meat & Poultry*** | | | | | | | | | | | | | |
| --- | --- | --- | --- | --- | --- | --- | --- | --- | --- | --- | --- | --- | --- | --- |
|  | | ***7α-OH*** | ***7β-OH*** | ***4β-OH*** | ***5,6α-Epoxy*** | ***5,6β-Epoxy*** | ***7-Keto*** | ***Triol*** | ***6-Keto*** | ***20α-OH*** | ***22-OH*** | ***24-OH*** | ***25-OH*** | ***Total COPs*** |
|  | ***(mg/100 g fat) ± STD*** | | | | | | | | | | | | | |
| ***MP1-RTE*** | | 5.95 ± 5.43 | 4.83 ± 4.30 | - | 0.10 ± 0.064 | 0.11 | 1.93 ± 1.57 | 0.15 ± 0.076 | 0.065 ± 0.029 | - | 0.068 ± 0.042 | - | 0.39 ± 0.32 | 15.27 ± 12.80 |
| ***MP2-RTE*** | | 4.20 ± 1.20 | 3.76 ± 1.06 | - | 0.12 ± 0.026 | 0.13 ± 0.040 | 1.14 ± 0.41 | 0.058 ± 0.0071 | 0.052 ± 0.020 | - | - | - | 0.36 ± 0.12 | 10.25 ± 2.80 |
| **MP3-RTE** | | 0.081 ± 0.019 | 0.060 ± 0.027 | - | - | - | 0.015 ± 0.0084 | - | - | - | - | - | - | 0.25 ± 0.027 |
| **MP4-RTE** | | 0.96 ± 0.14 | 1.13 ± 0.19 | - | - | - | 0.29 ± 0.021 | 0.22 ± 0.072 | - | - | 0.10 ± 0.034 | - | 0.14 ± 0.095 | 6.99 ± 1.85 |
| **MP5-RTE** | | 0.42 ± 0.11 | 0.32 ± 0.024 | - | - | - | 0.16 ± 0.099 | 0.082 ± 0.012 | 0.079 ± 0.020 | - | 0.17 ± 0.0081 | 0.050 ± 0.031 | 0.11 ± 0.021 | 2.17 ± 0.22 |
| **MP6-RTE** | | 15.85 ± 3.91 | 16.07 ± 4.20 | - | 0.26 ± 0.12 | 0.41 ± 0.13 | 3.75 ± 0.68 | 2.33 ± 0.37 | - | - | - | 0.40 ± 0.084 | - | 43.12 ± 8.98 |
| **MP7-RTE** | | 0.14 ± 0.029 | 0.15 ± 0.060 | - | - | - | 0.066 ± 0.044 | - | - | - | - | - | - | 0.36 ± 0.13 |
| **MP8-RTE** | | 0.54 ± 0.062 | 0.47 ± 0.066 | - | - | 0.13 ± 0.022 | 0.12 ± 0.039 | 0.053 ± 0.020 | - | - | 0.66 ± 0.043 | - | 0.19 ± 0.0078 | 3.09 ± 0.16 |
| **MP9-RTE** | | 0.27 ± 0.067 | 0.20 ± 0.070 | - | - | - | 0.085 ± 0.025 | - | - | - | - | - | - | 1.36 ± 0.38 |
|  | ***Seafood*** | | | | | | | | | | | | | |
|  | | ***7α-OH*** | ***7β-OH*** | ***4β-OH*** | ***5,6α-Epoxy*** | ***5,6β-Epoxy*** | ***7-Keto*** | ***Triol*** | ***6-Keto*** | ***20α-OH*** | ***22-OH*** | ***24-OH*** | ***25-OH*** | ***Total COPs*** |
|  | ***(mg/100 g fat) ± STD*** | | | | | | | | | | | | | |
| **S1-RTE** | | 0.052 ± 0.020 | 0.060 ± 0.013 | - | - | - | - | - | - | - | 0.090 ± 0.013 | - | - | 0.25 ± 0.026 |

|  | | ***Eggs & egg’s derivatives*** | | | | | | | | | | | | |
| --- | --- | --- | --- | --- | --- | --- | --- | --- | --- | --- | --- | --- | --- | --- |
|  | ***7α-OH*** | | ***7β-OH*** | ***4β-OH*** | ***5,6α-Epoxy*** | ***5,6β-Epoxy*** | ***7-Keto*** | ***Triol*** | ***6-Keto*** | ***20α-OH*** | ***22-OH*** | ***24-OH*** | ***25-OH*** | ***Total COPs*** |
|  | | ***(mg/100 g fat) ± STD*** | | | | | | | | | | | | |
| **E1-RTE** | 0.11 ± 0.051 | | 0.13 ± 0.024 | - | - | - | 0.062 ± 0.021 | - | - | 0.025 ± 0.017 | 0.069 ± 0.077 | - | 0.22 ± 0.036 | 0.91 ± 0.15 |
| **E2-RTE** | 0.47 ± 0.27 | | 0.32 ± 0.19 | - | - | - | 0.18 ± 0.088 | - | - | - | - | - | - | 1.35 ± 0.60 |
|  | | ***Baby Food*** | | | | | | | | | | | | |
|  | ***7α-OH*** | | ***7β-OH*** | ***4β-OH*** | ***5,6α-Epoxy*** | ***5,6β-Epoxy*** | ***7-Keto*** | ***Triol*** | ***6-Keto*** | ***20α-OH*** | ***22-OH*** | ***24-OH*** | ***25-OH*** | ***Total COPs*** |
|  | | ***(mg/100 g fat) ± STD*** | | | | | | | | | | | | |
| ***BF1-RTE*** | 8.97 ± 5.42 | | 24.61 ± 15.21 | - | 0.69 ± 0.36 | 1.39 ± 0.81 | 16.69 ± 5.45 | 2.63 ± 0.46 | - | - | - | - | 3.60 ± 1.22 | 62.55 ± 25.55 |
| ***BF2-RTE*** | 2.92 ± 0.73 | | 2.27 ± 0.64 | - | 0.044 ± 0.012 | 0.033 ± 0.0062 | 0.84 ± 0.34 | 0.22 ± 0.031 | 0.11 ± 0.064 | - | - | - | 0.24 ± 0.055 | 8.48 ± 2.46 |
| **BF3-RTE** | 0.10 ± 0.031 | | 0.065 ± 0.037 | - | - | - | - | - | - | 0.65 ± 0.17 | 0.33 ± 0.11 | 0.18 ± 0.47 | - | 2.65 ± 0.46 |
| **BF4-RTE** | 1.80 ± 0.22 | | 1.61 ± 0.24 | - | - | - | 0.57 ± 0.065 | 0.27 ± 0.086 | - | - | 0.20 ± 0.044 | - | 0.16 ± 0.056 | 7.32 ± 1.08 |
| **BF5-RTE** | 2.10 ± 0.50 | | 1.94 ± 0.37 | - | 0.052 ± 0.021 | 0.045 ± 0.027 | 0.59 ± 0.18 | 0.31 ± 0.091 | - | - | 0.10 ± 0.034 | - | 0.27 ± 0.16 | 6.44 ± 1.23 |
| **BF6-RTE** | 3.73 ± 1.98 | | 0.66 ± 0.12 | - | - | - | 0.67 ± 0.42 | 0.20 ± 0.034 | - | - | 0.077 ± 0.031 | - | - | 8.04 ± 3.16 |
| **BF7-RTE** | 2.12 ± 0.16 | | 2.38 ± 0.15 | - | - | - | 0.48 ± 0.15 | 0.39 ± 0.056 | - | - | 0.17 ± 0.069 | - | - | 6.13 ± 0.30 |
| **BF8-RTE** | 5.85 ± 1.82 | | 4.91 ± 1.12 | - | 0.030 ± 0.019 | 0.073 ± 0.039 | 1.68 ± 0.61 | 0.27 ± 0.10 | 0.12 ± 0.018 | - | - | - | 0.67 ± 0.16 | 14.02 ± 3.97 |
| **BF9-RTE** | 0.68 ± 0.12 | | 0.59 ± 0.17 | - | - | - | 0.24 ± 0.075 | 0.21 ± 0.050 | - | - | - | - | - | 8.14 ± 5.70 |
| ***BF10-RTE*** | 2.08 ± 0.63 | | 1.62 ± 0.46 | - | 0.027 ± 0.016 | 0.042 ± 0.032 | 0.65 ± 0.087 | 0.35 ± 0.041 | - | - | 0.067 ± 0.015 | - | 0.27 ± 0.047 | 6.36 ± 1.36 |
| ***BF11-RTE*** | 0.66 ± 0.092 | | 0.58 ± 0.020 | - | - | 1.05 ± 0.12 | - | 0.12 ± 0.036 | - | - | 2.48 ± 1.45 | - | - | 5.28 ± 1.49 |
| ***BF12-RTE*** | 0.41 ± 0.10 | | 0.40 ± 0.078 | - | - | - | 0.18 ± 0.013 | 0.10 ± 0.044 | - | - | 0.18 ± 0.11 | - | - | 2.05 ± 0.31 |
| ***BF13-RTE*** | 4.22 ± 3.77 | | 1.27 ± 0.89 | - | 0.062 ± 0.023 | 0.10 ± 0.025 | 1.31 ± 0.83 | 0.15 ± 0.03 | - | - | 0.12 ± 0.017 | 0.14 ± 0.029 | 0.47 ± 0.32 | 11.58 ± 7.07 |
|  | | ***Others*** | | | | | | | | | | | | |
|  | ***7α-OH*** | | ***7β-OH*** | ***4β-OH*** | ***5,6α-Epoxy*** | ***5,6β-Epoxy*** | ***7-Keto*** | ***Triol*** | ***6-Keto*** | ***20α-OH*** | ***22-OH*** | ***24-OH*** | ***25-OH*** | ***Total COPs*** |
|  | | ***(mg/100 g fat) ± STD*** | | | | | | | | | | | | |
| ***O1-RTE*** | 0.12 ± 0.018 | | 0.094 ± 0.023 | - | - | - | - | 0.053 ± 0.014 | 0.023 ± 0.011 | - | 0.099 ± 0.027 | - | - | 1.56 ± 0.28 |
| ***O2-RTE*** | 0.11 ± 0.037 | | 0.089 ± 0.015 | - | - | - | 0.035 ± 0.0079 | 0.060 ± 0.016 | - | 0.031 ± 0.020 | - | - | - | 0.61 ± 0.12 |
| ***O3-RTE*** | 0.57 ± 0.11 | | 0.60 ± 0.14 | - | 0.10 ± 0.030 | 0.055 ± 0.043 | 0.20 ± 0.060 | 0.13 ± 0.055 | - | - | 0.083 ± 0.0022 | - | - | 5.60 ± 1.28 |
| ***O4-RTE*** | 1.19 ± 0.78 | | 1.22 ± 0.79 | - | 0.066 ± 0.028 | 0.054 ± 0.053 | 0.30 ± 0.095 | 0.17 ± 0.063 | 0.078 ± 0.020 | - | 0.051 ± 0.027 | - | 0.15 ± 0.10 | 9.58 ± 2.73 |
