## Supplemental Table 5 for "Oxidative Status of the Ultra-Processed Foods in the Western Diet"

**Supplemental material**

**Table S5:** DOxS content in Fast Food group, expressed in mg per 100 g of fat.

|  | | ***Meat & Poultry*** | | | | | | | | | | | | |
| --- | --- | --- | --- | --- | --- | --- | --- | --- | --- | --- | --- | --- | --- | --- |
|  | ***7α-OH*** | | ***7β-OH*** | ***4β-OH*** | ***5,6α-E*** | ***5,6β-E*** | ***7-Keto*** | ***Triol*** | ***6-Keto*** | ***20α-OH*** | ***22-OH*** | ***24-OH*** | ***25-OH*** | ***Total COPs*** |
|  | | ***(mg/100 g fat) ± STD*** | | | | | | | | | | | | |
| **MP10** | 3.06 ± 1.37 | | 2.28 ± 1.21 | - | 0.051 ± 0.0052 | 0.068 ± 0.024 | 0.92 ± 0.48 | 0.12 ± 0.057 | - | - | - | 0.14 ± 0.094 | 0.21 ± 0.12 | 8.64 ± 3.04 |
| **MP11** | 0.31 ± 0.031 | | 0.29 ± 0.048 | - | - | - | 0.12 ± 0.052 | 0.033 ± 0.015 | 0.045 ± 0.034 | 0.11 ± 0.060 | 0.21 ± 0.17 | - | 0.17 ± 0.013 | 1.91 ± 0.084 |
| **MP12** | 1.00 ± 0.29 | | 0.72 ± 0.28 | - | - | - | 0.35 ± 0.083 | 0.030 ± 0.020 | 0.050 ± 0.0052 | - | - | 0.10 ± 0.024 | 0.11 ± 0.044 | 3.18 ± 0.65 |
| **MP13** | 0.26 ± 0.023 | | 0.28 ± 0.041 | - | - | - | 0.077 ± 0.16 | 0.024 ± 0.012 | - | - | - | - | 0.12 ± 0.036 | 1.67 ± 0.20 |
| **MP14** | 0.75 ± 0.044 | | 0.55 ± 0.086 | - | 0.056 ± 0.038 | 0.13 ± 0.041 | 0.54 ± 0.21 | 0.21 ± 0.037 | 0.070 ± 0.021 | 0.13 ± 0.036 | 0.20 ± 0.030 | - | 0.22 ± 0.038 | 14.64 ± 11.61 |
| **MP15** | 1.89 ± 0.66 | | 0.78 ± 0.10 | 0.49 ± 0.030 | 0.061 ± 0.013 | 0.11 ± 0.016 | 0.86 ± 0.33 | 0.12 ± 0.040 | - | - | - | - | - | 5.02 ± 1.08 |
| **MP16** | 0.69 ± 0.20 | | 0.75 ± 0.22 | - | 0.056 ± 0.037 | 0.044 ± 0.023 | 0.20 ± 0.033 | 0.053 ± 0.017 | - | 0.050 ± 0.017 | - | - | - | 3.90 ± 1.00 |
| **MP17** | 0.19 ± 0.44 | | 0.19 ± 0.037 | - | - | - | 0.63 ± 0.018 | 0.031 ± 0.018 | - | - | - | - | - | 1.55 ± 0.38 |
| **MP18** | 3.04 ± 0.32 | | 0.99 ± 0.42 | - | 0.023 ± 0.011 | 0.029 ± 0.0099 | 0.57 ± 0.089 | 0.067 ± 0.023 | 0.039 ± 0.014 | - | - | - | - | 5.72 ± 0.99 |
| **MP19** | 0.48 ± 0.044 | | 0.34 ± 0.036 | - | 0.030 ± 0.020 | 0.028 ± 0.0057 | 0.20 ± 0.040 | 0.11 ± 0.026 | 0.021 ± 0.0069 | - | 0.10 ± 0.032 | - | 0.049 ± 0.029 | 1.75 ± 0.077 |
| **MP20** | 0.18 ± 0.060 | | 0.16 ± 0.040 | - | - | - | 0.057 ± 0.035 | 0.037 ± 0.031 | - | - | - | - | 0.090 ± 0.055 | 0.91 ± 0.11 |
| **MP21** | 4.01 ± 0.081 | | 3.50 ± 0.14 | - | 0.060 ± 0.019 | 0.12 ± 0.041 | 1.42 ± 0.24 | 0.11 ± 0.038 | 0.069 ± 0.052 | 0.019 ± 0.0073 | 0.019 ± 0.016 | 0.18 ± 0.029 | 0.12 ± 0.0099 | 10.40 ± 0.71 |
| **MP22** | 1.81 ± 0.23 | | 2.61 ± 0.29 | - | - | - | 0.71 ± 0.034 | 0.071 ± 0.019 | - | - | - | - | - | 5.40 ± 0.56 |
| **MP23** | 16.80 ± 9.61 | | 10.23 ± 4.14 | 0.41 0.22 | 0.21 ± 0.078 | 0.52 ± 0.28 | - | 0.60 ± 0.32 | 0.22 ± 0.081 | 0.16 ± 0.045 | 0.43 ± 0.11 | - | 1.29 ± 0.64 | 39.13 ± 18.94 |
| **MP24** | 0.14 ± 0.036 | | 0.18 ± 0.035 | - | - | - | - | 0.036 ± 0.012 | - | - | - | - | 0.15 ± 0.064 | 2.53 ± 0.33 |
|  | | ***Seafood*** | | | | | | | | | | | | |
|  | ***7α-OH*** | | ***7β-OH*** | ***4β-OH*** | ***5,6α-Epoxy*** | ***5,6β-Epoxy*** | ***7-Keto*** | ***Triol*** | ***6-Keto*** | ***20α-OH*** | ***22-OH*** | ***24-OH*** | ***25-OH*** | ***Total COPs*** |
| ***(mg/100 g fat) ± STD*** | | | | | | | | | | | | | | |
| **S2** | 0.33 ± 0.075 | | 0.23 ± 0.029 | - | - | - | 0.15 ± 0.012 | 0.092 ± 0.029 | - | - | - | - | 0.16 ± 0.079 | 2.66 ± 0.44 |
| **S3** | 1.37 ± 0.83 | | 1.27 ± 0.72 | - | 0.055 ± 0.040 | 0.18 ± 0.037 | 0.43 ± 0.25 | 0.12 ± 0.069 | 0.081 ± 0.034 | 0.12 ± 0.065 | 0.20 ± 0.025 | - | 0.17 ± 0.042 | 7.27 ± 2.85 |
|  | | ***Others*** | | | | | | | | | | | | |
|  | ***7α-OH*** | | ***7β-OH*** | ***4β-OH*** | ***5,6α-Epoxy*** | ***5,6β-Epoxy*** | ***7-Keto*** | ***Triol*** | ***6-Keto*** | ***20α-OH*** | ***22-OH*** | ***24-OH*** | ***25-OH*** | ***Total COPs*** |
| ***(mg/100 g fat) ± STD*** | | | | | | | | | | | | | | |
| **O5** | 0.096 ± 0.036 | | 0.093 ± 0.033 | 0.031 ± 0.0090 | 0.021 ± 0.0041 | - | 0.084 ± 0.028 | - | - | - | - | 0.15 ± 0.039 | - | 3.16 ± 1.11 |
| **O6** | 0.52 ± 0.31 | | 0.30 ± 0.13 | - | - | - | 0.26 ± 0.18 | 0.10 ± 0.017 | - | 0.16 ± 0.10 | 0.18 ± 0.017 | - | tr | 1.97 ± 0.61 |
| **O7** | 0.81 ± 0.038 | | 0.60 ± 0.044 | - | 0.068 ± 0.036 | 0.080 ± 0.018 | 0.22 ± 0.021 | 0.075 ± 0.015 | - | 0.11 ± 0.042 | - | - | - | 2.40 ± 0.14 |
| **O8** | 0.11 ± 0.073 | | 0.096 ± 0.045 | - | - | - | - | 0.021 ± 0.013 | - | - | 0.024 ± 0.019 | - | 0.16 ± 0.033 | 3.14 ± 1.56 |
| **O9** | 0.099 ± 0.037 | | 0.082 ± 0.033 | - | - | - | 0.056 ± 0.027 | 0.033 ± 0.0065 | - | 0.052 ± 0.013 | - | - | 0.19 ± 0.049 | 9.62 ± 2.24 |
| **O10** | 0.42 ± 0.052 | | 0.37 ± 0.060 | - | - | 0.10 ± 0.022 | - | 0.041 ± 0.017 | - | - | 0.52 ± 0.030 | - | 0.15 ± 0.011 | 2.33 ± 0.21 |

- NO Detected, FF with a blue background, RTE with green background.
