## Supplemental Table 6 for "Oxidative Status of the Ultra-Processed Foods in the Western Diet"

**Table S6**: Potential biomarkers per UPFs group and category (with MetaboAnalyst).

|  |  | **Biomarker** | **AUC** |
| --- | --- | --- | --- |
| ***Groups*** | RTE | Brassicasterol | 0.78 |
|  | FF | 7α-OH and 7β-OH | 0.82 and 0.80 |
| ***Categories*** | Dairy | Brassicasterol | 0.73 |
|  | Eggs & derivatives | Stigmasterol and β-Sitosterol | 0.87 and 0.80 |
|  | Meat & poultry | 7α-OH | 0.75 |
|  | Seafood | 5,6β-Epoxycholesterol | 0.43 |
|  | Baby food | β-Sitosterol | 0.64 |
|  | Others | Campesterol | 0.72 |
